# A systematic review and meta-analysis of the risk of hepatitis B virus (HBV) genotypic resistance in people treated with entecavir or tenofovir

**DOI:** 10.1101/2023.11.08.23298154

**Authors:** Sheila F Lumley, Marion Delphin, Jolynne F Mokaya, Cedric CS Tan, Emily Martyn, Motswedi Anderson, Ka Chun Li, Elizabeth Waddilove, Gloria Sukali, Louise O Downs, Khadija Said, Dorcas Okanda, Cori Campbell, Eli Harriss, Yusuke Shimakawa, Philippa C Matthews

## Abstract

**Background:** As nucleos/tide analogue (NA) therapy for chronic Hepatitis B virus (HBV) infection becomes more widely indicated and available, understanding drug resistance is essential. We performed a systematic review and meta-analysis to estimate the risk of genotypic resistance to tenofovir and entecavir.

**Methods:** We searched nine databases up to 29-Aug-23. We included studies of HBV infection featuring >10 individuals, written in English, reporting tenofovir or entecavir treatment ≥48 weeks, with assessment of HBV genotypic resistance. Data were analysed according to prior exposure history to NA, and treatment with tenofovir or entecavir. Analyses were performed in R.

**Results:** 62 studies involving a total of 12,358 participants were included. For tenofovir, pooled resistance risk was 0.0% at all time points, whether previously NA naive (11 studies; 3778 individuals) or experienced (19 studies; 2059 individuals). For entecavir, in treatment-naive individuals (22 studies; 4326 individuals), risk of resistance increased over time to 0.9% at ≥5 years (95%CI 0.1-2.3%). Entecavir resistance was increased in NA-experienced individuals (18 studies;1112 individuals), to 20.1% (95%CI 1.6-50.1%) at ≥5 years. There was a lack of consistent definitions, poor global representation and insufficient metadata to support subgroup analysis.

**Discussion:** Based on existing data, tenofovir has an excellent resistance profile. More resistance is seen with entecavir, particularly in treatment-experienced groups. Due to data gaps, we may have under-estimated the true risk of resistance. Robust prospective data collection is crucial as treatment is rolled out more widely.

## Introduction

Chronic hepatitis B virus (HBV) infection (CHB) is a global public health challenge, affecting an estimated 300 million people worldwide, despite the availability of effective vaccines ^1^. CHB continues to be a significant cause of liver-related morbidity and mortality, accounting for up to one million deaths a year. The management of CHB centres on antiviral therapy, with the primary goal of suppressing viral replication, reducing liver inflammation, and preventing progression to cirrhosis and hepatocellular carcinoma. Lowering HBV viral load (VL) also reduces the risk of transmission; this is best exemplified by the use of prophylaxis for pregnant women at the highest risk of mother-to-child-transmission ^2^.

Nucleoside analogues (e.g. entecavir (ETV)) and nucleotide analogues (e.g. tenofovir (TFV), administered as tenofovir disoproxil (TDF) or tenofovir alafenamide (TAF)) (NAs) are first line treatments^3,4^, while lamivudine (LAM) and adefovir (ADV) have been largely phased out due to the predictable selection of viral resistance-associated polymorphisms (RAMs) ^5^. LAM and TFV were adopted from use in HIV therapy due to the structural similarity of the HIV and HBV reverse transcriptases (RT).

Although the emergence of resistance to TFV and ETV is thought to be infrequent ^5,6^, assimilating evidence for its risk and impact is crucial to determine the extent to which it represents a potential challenge. Prior exposure to NA can influence subsequent development of drug resistance due to the presence of shared RAMs (Table 1A). Although ETV has a high genetic barrier to resistance, resistance has been reported, especially in patients with genotypic resistance to LAM ^7–10^. The extent to which TFV resistance is a real-world problem - either for individual patients or on public health grounds - remains uncertain and is likely to vary between population settings. Individuals have been identified with virological breakthrough despite confirmed adherence to TDF, however few RAMs have been consistently validated in vivo and in vitro ^11–13^, it is likely that combinations of multiple polymorphisms are required to produce clinically significant resistance ^6,11,12,14^.

**Table 1:** Antiviral drug resistance mutations in HBV. **Table 1A:** Antiviral drug resistance mutations and cross-resistance in chronic HBV, as defined in EASL guidance^6^

| HBV variant | LAM | LDT | ETV | ADV | TDF/TAF* |
| --- | --- | --- | --- | --- | --- |
| Wild-type | S | S | S | S | S |
| M204V | R | S | I | I | S |
| M204I | R | R | I | I | S |
| L180M + M204V | R | R | I | I | S |
| A181T/V | I | I | S | R | I |
| N236T | S | S | S | R | I |
| L180M + M204V/I<br>+/- I169T +/- V173L +/- M250V | R | R | R | S | S |
| L180M + M204V/I<br>+/- T184G +/- S202I/G | R | R | R | S | S |

The amino acid substitution profiles are shown in the left column and the level of susceptibility is given for each drug: S (sensitive), I (intermediate/reduced susceptibility), R (resistant).
ETV, Entecavir; TDF Tenofovir disoproxil fumarate; TAF, Tenofovir alafenamide; LAM, lamivudine; ADV, adefovir
\*In vitro data for Tenofovir, in vivo data for TDF, no clinical data for TAF.

The landscape of HBV treatment is changing; the World Health Organization (WHO) and other bodies responsible for clinical recommendations are reviewing and revising guidelines, with a view to simplifying and expanding treatment eligibility ^15,16^. As therapy becomes more widely available, there is a pressing need to consider drug resistance as a potential barrier to its success, especially as most patients require long-term therapy to achieve sustained virological suppression. Since NAs are not curative, new drug agents are in development (e.g. small interfering RNA (siRNA) in phase II/III studies ^17^), but many of these first require suppression of HBV on first line NA therapy, so even as novel treatments become available, NA agents may still represent a first-line foundation. In addition, as NA drugs are safe, cheap and widely available, these will remain a practical first-line option for many population settings prior to the universal roll out of new alternatives.

The primary objective of this systematic review and meta-analysis is to summarise the risk of drug resistance to ETV and TFV over time in people receiving treatment for CHB. Our review aims to inform clinical practice, global treatment guidelines, provision of laboratory infrastructure and careful data collection, ultimately improving care and clinical outcomes, and informing progress towards global elimination goals for HBV infection.

## Methods

We set out to summarise the risk of HBV genotypic drug resistance in people living with CHB exposed to ETV or TFV for 48 weeks or longer. We conducted a systematic review and meta-analysis using the Preferred Reporting Items for Systematic Review and Meta-analysis (PRISMA) guidelines; a PRISMA checklist is provided in supplementary table 1. The review protocol can be found under PROSPERO registration number CRD42023424125. Further methodological details can be found in the supplementary text 1 and 2.

### Definitions of drug resistance

We used standardised nomenclature for clinical, phenotypic and genotypic resistance to NA therapy throughout ^11^.

‘Clinical resistance’ consists of individuals with:

i. Virologic breakthrough (VBT) (an increase in serum HBV DNA by ≥1 log10 IU/mL above nadir on ≥2 occasions 1 month apart, in a treatment-compliant patient);
ii. Primary non-response (inability of NA treatment to reduce serum HBV DNA by ≥1 log10 IU/mL after the first 6 months of treatment); or
iii. Partial response (detectable HBV DNA during continuous therapy).

‘Phenotypic resistance’ is defined as the ‘decreased susceptibility of an HBV polymerase to an antiviral treatment in vitro’ ^11^.

‘Genotypic resistance’ is defined as viral populations bearing amino acid substitutions in RT that have been shown to confer resistance to antiviral drugs in phenotypic assays. These mutations are usually detected in individuals with VBT but may also be present in those with persistent viremia ^11^ (Table 1A, 1B). The accepted standard for characterising the sequence of the polymerase domain of RT in order to identify mutations is sequencing of the PCR amplified product ^11^.

**Table 1B:** Additional putative TFV RAMs not listed in EASL guidance ^13^.

**1B: Additional putative TFV RAMs not listed in EASL guidance<sup>13</sup>**
| Putative RAM mutation/<br>combination of RAMs |  | Evidence |
| --- | --- | --- |
| A194T |  | Conflicting evidence<sup>29-32</sup>. |
| S106C + H126Y + D134E +/- L269I |  | Conflicting evidence<sup>14,31</sup> |
| L180M +<br>M204I/V | + R153W/Q + I163V | <sup>33</sup> |
|  | + V173L | Conflicting evidence<sup>33-36</sup>. |

We planned to validate the presence of drug resistance against a pre-specified list of mutations in the viral RT using a standardised numbering system ^18^. All the RAMs we sought to identify have been evidenced through phenotypic assays (Table 1). We accepted the conclusion presented in the primary paper regarding the presence or absence of genotypic resistance, as specific HBV mutations for each individual were not consistently listed in each paper, and sequences were typically not available in the public domain. For TFV, where there is less clarity regarding the definition of genotypic resistance, for each paper we summarised which RAMs were identified (Table 1A and 1B), and whether or not they were reported as resistant (Supplementary table 2).

### Data sources and search strategy

The following bibliographic databases and trial registries were initially searched by an information specialist (EH) on 01/06/2023 and updated 29/08/23 for studies published from database inception to the search date: Ovid MEDLINE, Ovid Embase, Ovid Global Health, Scopus, the Cochrane Central Register of Controlled Trials, clinicaltrials.gov, the ISRCTN Registry, and the WHO International Clinical Trials Registry. We searched the databases using relevant index terms and free text terms, synonyms, and phrases in the title and abstract fields for relevant papers to meet the aims of this review. The full strategies are available in supplementary text 1.

### Selection criteria

Inclusion criteria were:

i. study of ≥10 people living with CHB exposed to TFV and/or ETV monotherapy for ≥48 weeks, *and*
ii. study reports risk of clinical resistance for either TFV and/or ETV, *and*
iii. study reports genotypic resistance for either TFV and/or ETV, based on viral sequencing.

Exclusion criteria were:

i. not an original research article,
ii. incorrect study population (e.g. condition not CHB, individuals not taking TFV or ETV monotherapy, all individuals non-suppressed on TFV/ETV treatment, clinical phenotype not reported, unable to separate naive from experienced outcomes)
iii. ) sample size <10 individuals (within the synthesis group of interest (ie. NA naive vs. experienced, TFV vs. ETV)),
iv. treatment duration <48 weeks or not specified,
v. viral sequencing not used or HBV not sequenced after TFV/ETV treatment,
vi. not in English.

Criterion (ii) aimed to exclude studies that could potentially introduce selection bias into our estimate of risk, for example cohorts of individuals who all have VBT on TFV/ETV, or cohorts where all individuals have been referred for resistance testing.

### Screening and data extraction

Our team first undertook screening of titles and abstracts, then full text review for data extraction, with at least two reviewers independently screening at each stage (Supplementary text 2). Two review authors independently extracted information for each of the eligible studies after training and piloting the Covidence data extraction tool before use. The key outcome measure sought was the number of individuals developing genotypic resistance. The key effect measure was the cumulative incidence of genotypic resistance (defined above), calculated by dividing the number of individuals with genotypic resistance during a particular time period divided by the total number receiving treatment at the beginning of that time period.

Risk of bias was assessed independently by two reviewers using a modified five question Joanna Briggs Institute quality assessment tool (Supplementary text 2). Publication bias was assessed with funnel plots of study size against log odds (= ln(cases +0.5 / (sample size +0.5) - (cases+0.5))) ^19^, with the addition of 0.5 to cases and sample size to allow studies with zero events to be plotted.

### Data analysis

We analysed our data in four groups determined a priori:

i. “Naive/Tenofovir” - previously NA naive individuals treated with TFV,
ii. “Experienced/Tenofovir” - previously NA experienced individuals treated with TFV,
iii. “Naive/Entecavir” - previously NA naive individuals treated with ETV,
iv. “Experienced/Entecavir” - previously NA experienced individuals treated with ETV.

The primary objective was to estimate the risk of genotypic resistance and associated 95% confidence interval (CI) for all studies in the four groups detailed above after 1yr, 2yr, 3yr, 4yr and ≥5yr of TFV/ETV treatment. Time points were grouped as follows: 1 year (17 months or less), 2 years (18 - 29 months), 3 years (30 - 41 months), 4 years (42 - 53 months), ≥5 years (54 months and over). Where the same or overlapping datasets were presented in two studies categorised within the same time point, the report with the largest sample size was included. If the sample size was the same, the study with the lowest risk of bias was included. We produced forest plots and pooled risk across studies using a random-effects meta-analysis with a Freeman-Tukey double arcsine transformation as this was a meta-analysis of proportion data with a low frequency of events including multiple incidences of zero events.

Our secondary objective, if the number of studies allowed, was to estimate risk of antiviral resistance across subgroups (study design, WHO region, age, proportion of males, HIV coinfection status, HBeAg status, presence of baseline resistance mutations and criteria for sequencing), calculating the antiviral resistance risk and associated 95% CI.

We assessed heterogeneity arising from clinical and methodological diversity by performing sensitivity analyses where heterogeneity (I^2^) in the primary analysis was present (ie. where pooled estimate was generated and I^2^ ≠ 0%), comparing changes to I^2^. Sensitivity analyses planned were (i) restricting analysis to study size >30, (ii) excluding studies with high risk of bias, (iii) restricting to clinical trials, (iv) restricting to studies where all viraemic samples are sequenced (rather than only those meeting the definition for VBT) and (v) excluding those where all individuals were known to have genotypic resistance to another antiviral agent (e.g. LAM. ADV or ETV (for studies of TFV)) at baseline. We also qualitatively assessed outliers where the study 95% confidence interval lies outside the 95% confidence interval of the pooled effect, however only one study met this criterion, so in addition for studies of TFV we reviewed all studies reporting resistance and for studies of ETV we reviewed the highest and lowest estimates in groups where I^2^ ≠ 0%.

Statistical analyses were performed using the metaprop package in R (version 4.2.2).

## Results

### 1. Study characteristics

Among 7387 studies identified, 590 full texts were screened for eligibility and 62 (involving 12,358 participants) were included in our analyses (Figure 1). Of these, 11 studied NA naive individuals subsequently receiving TFV (Naive/Tenofovir), 22 studied NA experienced individuals treated with TFV (Experienced/Tenofovir), 24 studied NA naive individuals treated with ETV (Naive/Entecavir) and 18 studied NA experienced individuals treated with ETV (Experienced/Entecavir) (Figures 1, 2A, 2B).

**Figure 1.**
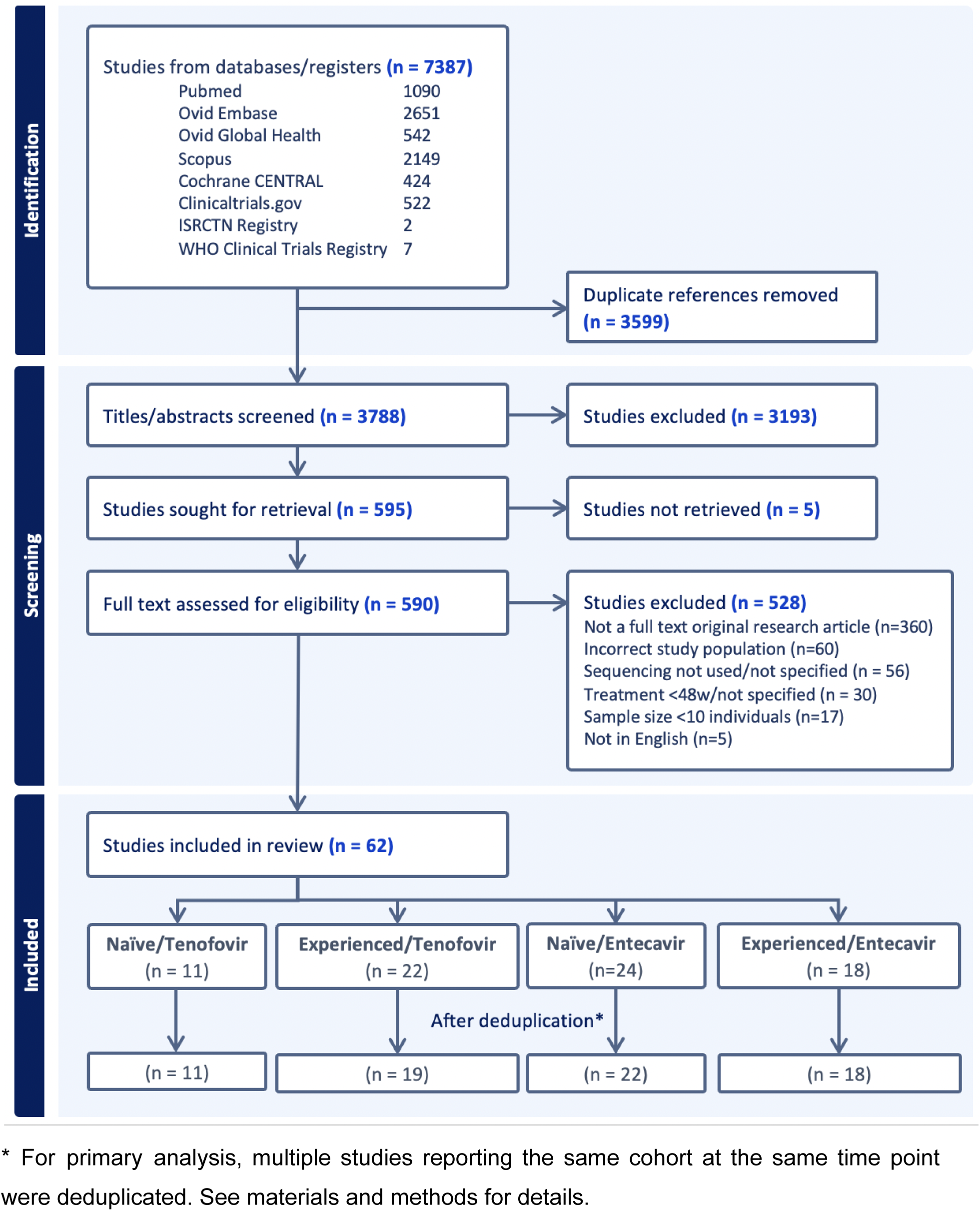
PRISMA flowchart. * For primary analysis, multiple studies reporting the same cohort at the same time point were deduplicated. See materials and methods for details.

**Figure 2.**
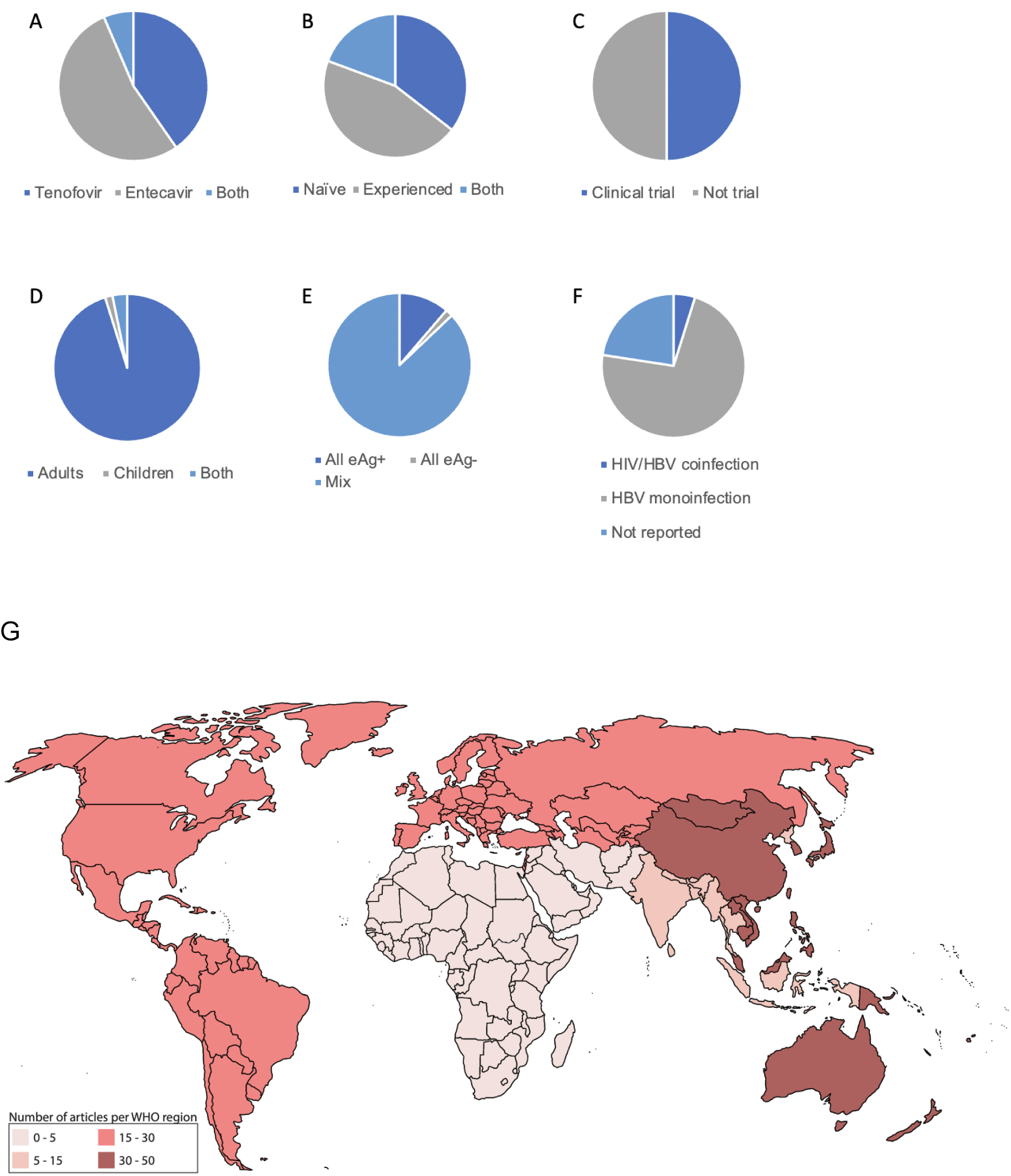
Characteristics of 62 studies included in our systematic review and meta analysis to determine the risk of TFV and ETV resistance in individuals receiving treatment for CHB infection. (A) NA being studied, (B) Prior exposure to NA agents, (C) Study type, (D) Population age category, (E) HBeAg status of participants at baseline, (F) Presence of HBV/HIV coinfection, (G) Distribution of studies per WHO regions.

Study characteristics are presented in Supplementary table 3 and Figure 2. There was an even split of study type; 50% of studies were clinical trials and 50% observational cohort studies (Figure 2C). The majority (59/62, 95%) were studies of adults, one studied children only and two included both (Figure 2D). Most (54/62, 87%) included a mix of HBeAg positive and negative individuals, one reported data from HBeAg negative individuals and seven from HBeAg positive individuals only (Figure 2E). HBV/HIV coinfection was present in two studies, however in general reporting was poor with HIV status not reported in 14/62 (22%) of studies (Figure 2F). The global distribution was skewed to the WHO Western Pacific region (49 studies), with only two studies in the WHO African region (Figure 2G).

### 2. Risk of TFV and ETV resistance over time

The primary objective was to estimate the risk of genotypic resistance in the four main treatment groups (based on prior exposure/current treatment) after 1, 2, 3, 4 and ≥5 years of treatment.

#### i) Resistance in previously NA naive individuals treated with TFV

Eleven studies including 3778 individuals reported risk of TFV resistance in the “naive/tenofovir” groups (Figure 3A, Supplementary table 3). Very low risk resistance to TFV was reported, with pooled estimates of 0.0% at all time points (95% CI 0.0% - 0.2% at 1 year, 95% CI 0.0 - 0.0% at 2 years and 95% CI 0.0 - 0.1% at ≥5 years).

**Figure 3.**
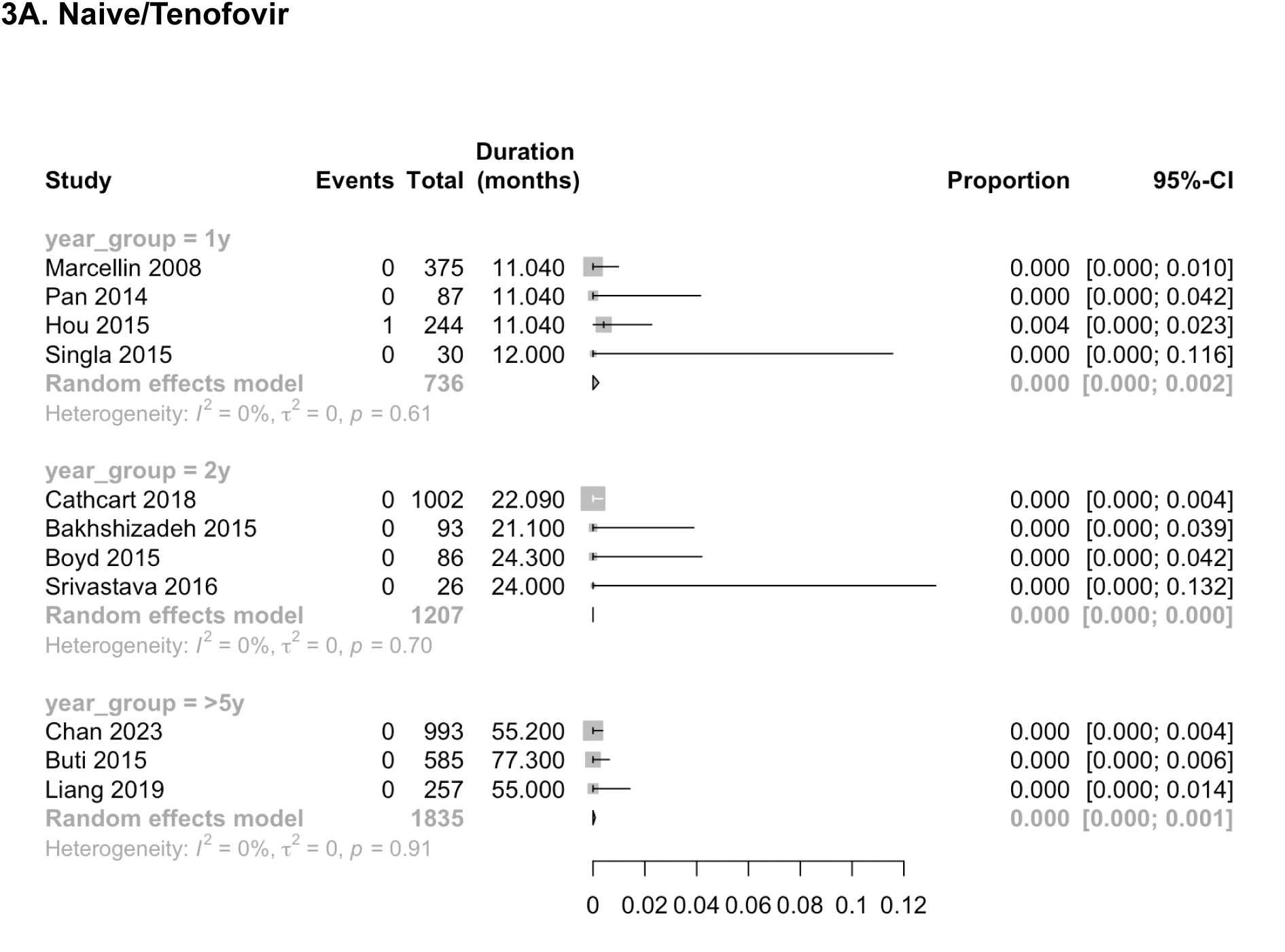
Meta-analysis of risk of TFV and ETV resistance over time. Forest plots for each synthesis, grouped by previous NA exposure, current NA treatment and duration of current NA treatment **Figure 3A. Naive/Tenofovir**

#### ii) Resistance in NA experienced individuals treated with TFV

Twenty two studies reported risk of TFV mutations in “experienced/tenofovir” groups. Three studies were excluded from meta-analysis as data from the same cohort were published multiple times within the same time point (see Methods for details of deduplication). The remaining 19 studies involved 2059 individuals (Figure 3B, Supplementary table 3). All pooled estimates were 0.0%, with 95% confidence intervals of 0.0 - 0.7% at 1 year, 0.0 - 0.6% at 2 years, 0.0 - 2.2% at 3 years and 0.0 - 0.0% at ≥5 years.

**Figure 3B.**
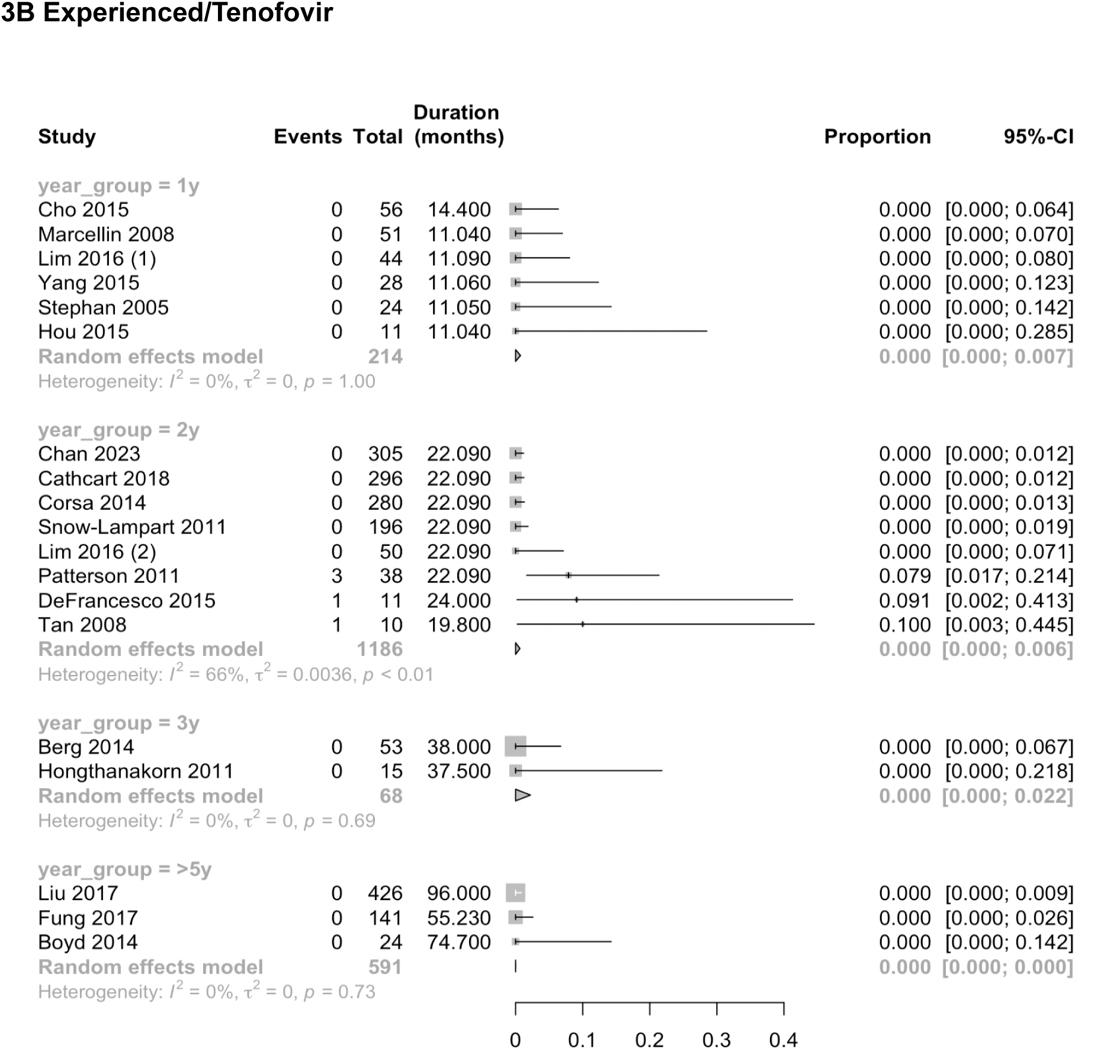
Experienced/Tenofovir

#### iii) Resistance in NA naive individuals treated with ETV

Twenty four studies reported risk of ETV resistance in “naive/entecavir: groups. Two studies were excluded as data from the same cohort was published multiple times within the same time point (see methods for deduplication details). The remaining 22 studies involved 4326 individuals (Figure 3C, Supplementary table 3). A low risk of resistance that increased with the duration of ETV treatment was seen. After 1 year on ETV, the pooled estimate of resistance was 0.0% (95% CI 0.0 - 0.2%), after 2 years 0.3% (95% CI 0.0 - 0.9%), after 4 years 0.6% (95% CI 0.00 - 0.27%) and at ≥5 years 0.9% (95% CI 0.1 - 2.3%).

**Figure 3C.**
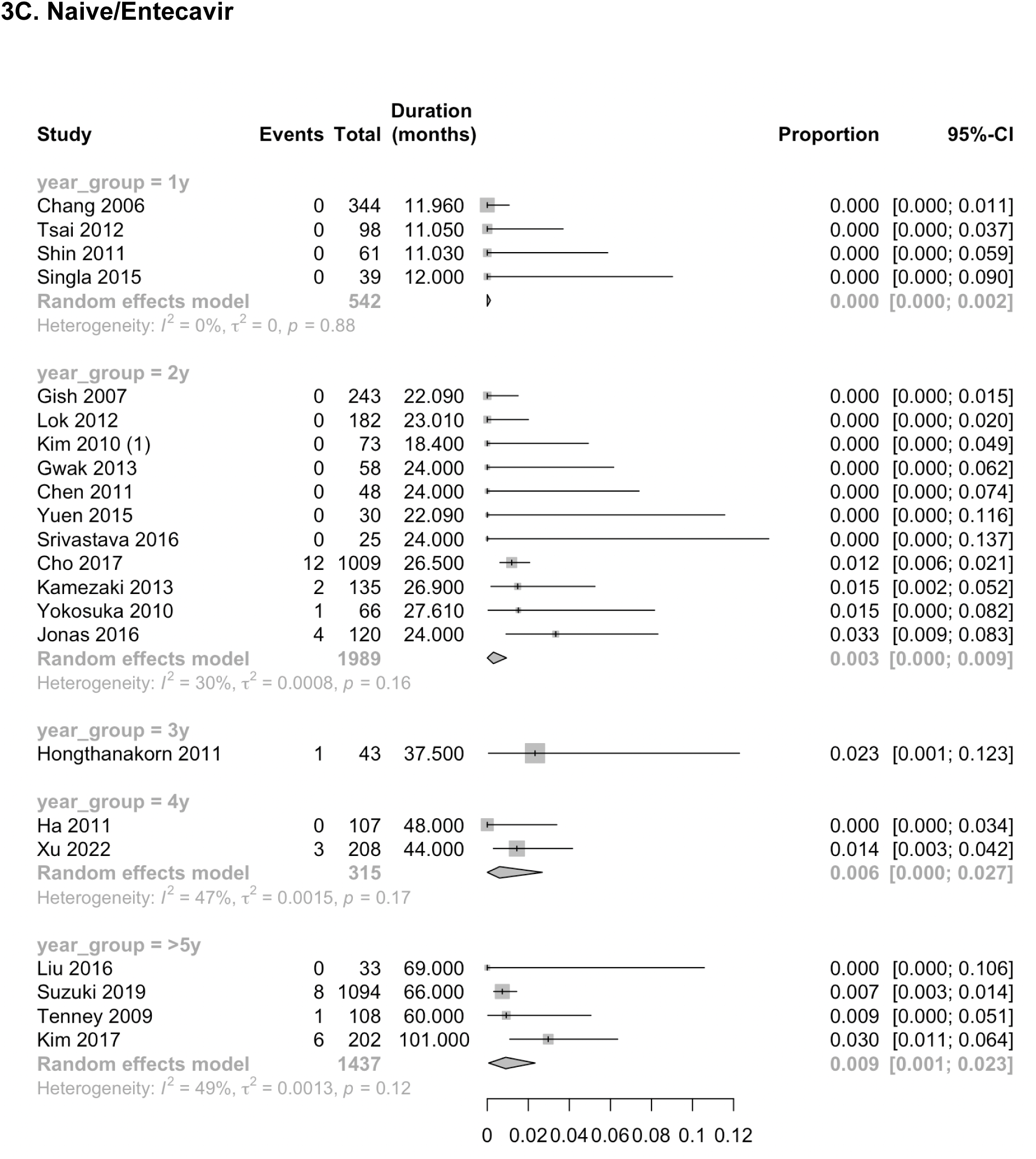
Naive/Entecavir

#### iv) Resistance in NA experienced individuals treated with ETV

Eighteen studies involving 1112 individuals reported risk of ETV mutations in “experienced/entecavir” groups (Figure 3D, Supplementary table 3). ETV resistance was common and increased over time. At 1 year the pooled estimate of resistance was 0.2% (95% CI 0.0 - 1.2%), at 2 years 17.0% (95% CI 5.4 - 32.9%), at 3 years 22.6% (8.2 - 41.0%) and ≥5 years 20.1% (1.6 - 50.1%).

**Figure 3D.**
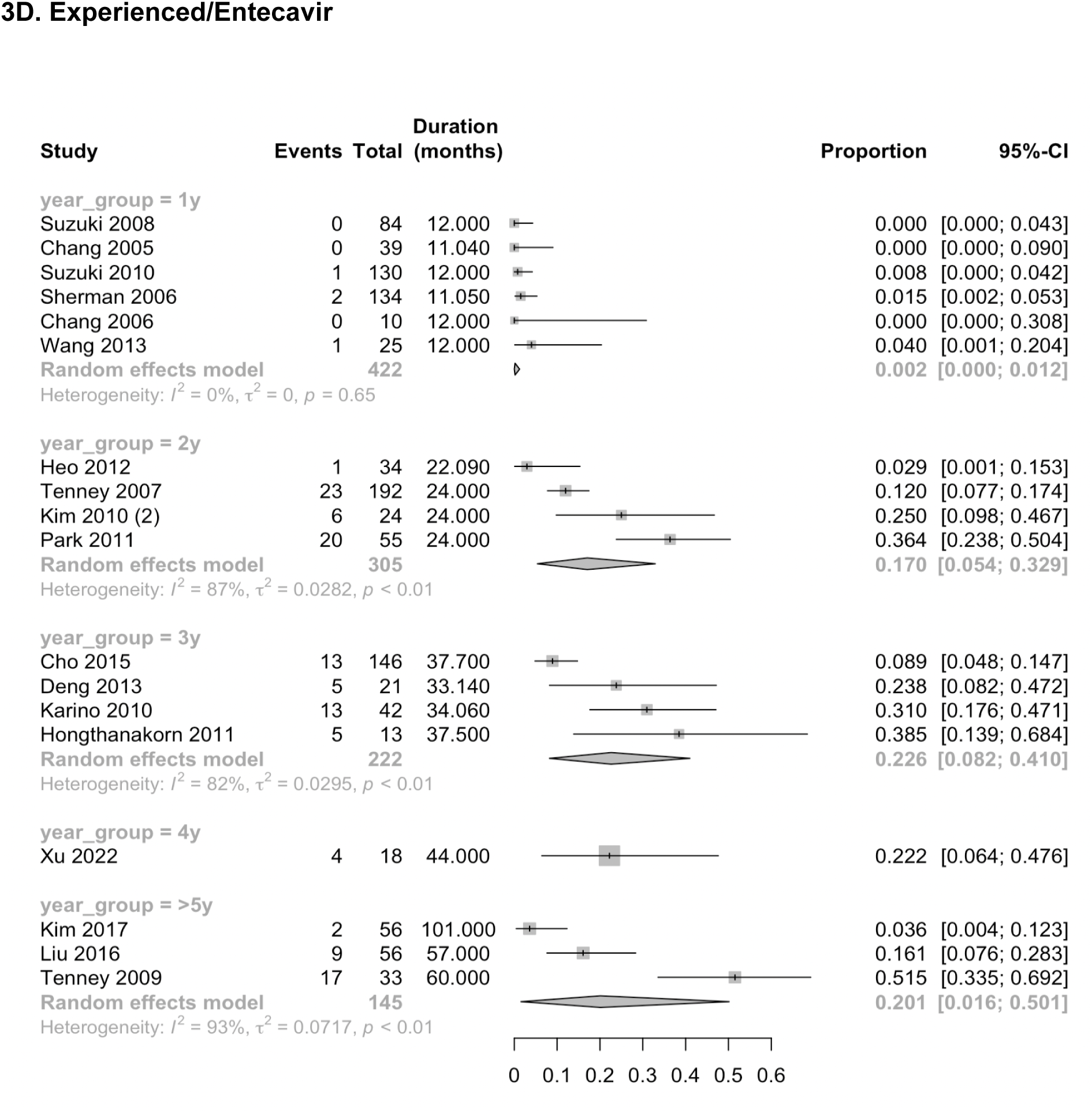
Experienced/Entecavir

### 3. Subgroup analysis

Due to the small number of studies within each exposure/treatment/year group category, and variable reporting of age/sex, subgroup analysis was not performed.

### 4. Sensitivity and outlier analysis

We explored study heterogeneity arising from clinical and methodological diversity by performing sensitivity analyses where heterogeneity in the primary analysis was present (ie. where pooled estimate was generated and I^2^ ≠ 0%) and also performed a qualitative assessment of outliers (see supplementary text 3 and supplementary table 4). We did not identify any variables consistently affecting the heterogeneity of estimates.

### 5. Quality and risk of bias

The quality/risk of bias scores for individual studies are presented in Supplementary table 3 and Supplementary figure 1. In the primary analysis, distribution of studies of low/moderate risk was consistent across the naive/tenofovir (5 low:6 moderate) experienced/tenofovir (8 low:10 moderate:1 high), naive/entecavir (10 low:12 moderate) subgroups. In the experienced/entecavir group, a higher proportion of moderate risk studies was seen (6 low:11 moderate:1 high).

Criteria for inclusion were well reported in most studies (87%) included in the primary analysis (Supplementary figure 1), however there was clinical heterogeneity between the populations included, particularly within the NA experienced cohorts. Inclusion criteria ranged from individuals with prior NA treatment without phenotypic/genotypic resistance, to populations where all had phenotypic but not genotypic resistance and others where all had phenotypic/genotypic resistance (with RAMs to other NA) before starting TFV/ETV. All studies used objective standard criteria for the diagnosis of CHB.

Study subjects were described in detail in 82% of the studies included in the primary analysis. However in some studies, demographic characteristics at baseline were not provided at the individual level, meaning subgroup analyses based on age and sex could not be performed. There was variable reporting of suspected risk factors for resistance e.g. HIV status, HBeAg status, drug adherence and genotype of infection (which is of importance, as some genotypes have certain RAMs as wild type consensus ^20^). The duration of TFV/ETV treatment spanned a wider range in cohort and cross-sectional studies compared to clinical trials where all individuals were followed up for the same duration, potentially providing a less accurate estimate of the relationship between duration of therapy and risk of resistance. There was sometimes a lack of clarity about prior agents used, duration of prior therapy and breaks in therapy. We were unable to consistently extract follow up data (Supplementary figure 2); for consistency, denominators were recorded at baseline, however in some cohorts some individuals switched treatment for undefined reasons, there was lack of clarity regarding which individuals had HBV sequencing undertaken on cessation/switching therapy, and in some cases individuals were lost to follow up.

Outcomes were measured in a valid way in 92% of studies. However, a range of sequencing methods were used (from those reporting consensus sequences to quasispecies), with varying sensitivity for detecting resistance. Criteria for sequencing differed between studies, some sequencing all viraemic individuals but others only sequencing those with VBT, risking underestimating the risk of resistance. Definitions of drug resistance were inconsistent between studies of TFV (Table 1, Supplementary table 2). In some papers, the criteria used to determine resistance were not specified, and when they were, studies used different thresholds/criteria to call resistance. Since not all polymorphisms were listed for each individual and the majority of sequences were not published, we had to use the authors’ judgement on what constituted genotypic resistance, rather than being able to take an objective view using the same criteria across all studies (ie. a predefined list of relevant RAM combinations). Furthermore, sequencing was not always successful, which may lead to an under-estimation of resistance risk. The relationship between clinical and methodological sources of heterogeneity and the observed risk of genotypic resistance over time is presented in Supplementary figure 3.

Due to the decision made a priori to present results of NA naive and NA experienced individuals separately, some papers were excluded as resistance risk was not reported for naive and experienced subgroups separately, or the denominator was not clear and resistance risk could not be calculated ^21–27^.

Publication bias was assessed with funnel plots of study size against log odds. For TFV, most studies reported zero events and therefore small-study effects could not be assessed. For ETV, the adapted funnel plot was asymmetrical for studies of individuals who were previously NA naive and symmetrical for studies of individuals who were previously NA experienced, indicating that there might be a publication bias for studies of NA naive individuals, towards smaller studies reporting a higher risk (Supplementary figure 4).

## Discussion

### Summary

As NA therapy is more widely rolled out as a component of interventions aiming to deliver global elimination targets for HBV infection by the year 2030, there is an imperative to collate and analyse data about outcomes on treatment, including groups in whom there is a risk of drug resistance. To our knowledge, this is the first systematic review and meta-analysis of TFV and ETV resistance. Based on these data we conclude that TFV has an excellent resistance profile in both treatment naive and experienced individuals, even with long term therapy, confirming its high genetic barrier to the development of resistance. There was a higher risk of genotypic resistance with ETV treatment, particularly in patients with prior failure (and/or confirmed resistance) to other NAs. This reaffirms the current protocols for use of TFV as the first line NA in patients with prior failure of or resistance to other agents ^3,5,6,28^. However, we recognise that most studies include individuals and groups at the highest risk of resistance, and our approach may therefore under-estimate resistance.

### Caveats and limitations

This meta-analysis was limited by the small number of trials within each group. Clinical and methodological heterogeneity was observed (see Quality section for discussion), with variation in inclusion criteria (including presence/absence of baseline RAMs) and varied definitions of TFV drug resistance potentially impacting on resistance risk estimates.

In order to optimise the quality and reduce bias, we excluded smaller studies and those only focusing on drug-resistant populations, and did not include studies that did not present viral sequence data. We recognise this focused approach means we have not accounted for the full spectrum of published data on drug resistance; assimilation of evidence from studies specifically focusing on drug resistant populations could be a future aspiration to collect data on risk factors, clinical characteristics and sequence motifs associated with resistance. Further investment and research is required to increase the sensitivity of sequencing methods to enable sequencing of lower viral load samples and to ensure methods are pan-genotypic. Future studies should aim to identify novel as well as known RAMs.

Half of the studies included were clinical trials, which are poorly representative of the general population, typically not recruiting (or subsequently excluding) key populations that may be at highest risk of drug resistance, for example those with incomplete adherence, HIV +/− HCV/HDV coinfection, multimorbidity, liver disease, substance abuse, highly mobile populations (including migrants, travellers, people experiencing homelessness) and those who are pregnant or breastfeeding. For these reasons, observed resistance rates within this dataset are likely to underestimate real world development of resistance when NA therapy is used at scale in the wider population. Whilst drug adherence is less likely to be an issue during clinical trials, it may be a risk factor for resistance, and itself has multiple drivers which include lack of education, socioeconomic deprivation, out-of-pocket costs to patients, stigma and discrimination.

Furthermore, there was poor global representation of studies, studies typically focused on well characterised populations in resource-rich settings, excluding populations with the highest burden of infection and associated liver disease. We note particular underrepresentation of the WHO Africa region (Figure 2G), a region with risk factors for high risk of resistance (poor/inconsistent access to treatment and where HIV/HBV are co-endemic with high exposure to antiretroviral therapy). This inequity and neglect has been recognised as an issue in HBV clinical trials more broadly (unpublished results from our team), and is a key knowledge gap when considering treatment approaches globally, relevant to global elimination goals.

### Conclusions and recommendations for the field

Undertaking a systematic analysis has provided insights into challenges in the current literature, and highlights important avenues for future scrutiny, with immediate translational implications as more people become eligible for, and can access, NA therapy. We can thus conclude with key aspirations for future studies (Table 2). Future studies should strive to address global representation, to generate high quality, long duration, real world data that includes populations with risk factors for resistance. Standardised reporting of drug resistance, publication of clinical metadata and sharing of sequence data are essential attributes of reporting. These data are essential to guide strategies for implementation of effective therapy that will support progress towards HBV elimination worldwide.

**Table 2:** Summary of recommendations to the field for clinical studies investigating HBV drug resistance.

| Domain | Recommendation |
| --- | --- |
| Study type | - 'Real world' prospective cohort studies<br>- Improved global distribution of studies, increase data for countries with high HBV prevalence<br>- Include populations typically excluded from clinical trials e.g. pregnant/breastfeeding, incomplete adherence, HIV +/- HCV/HDV coinfection, multimorbidity, liver disease, substance abuse, highly mobile populations (including migrants, travellers, people experiencing homelessness), children and older age.<br>- Long-term follow up over the lifetime of an individual<br>- Review of drug resistance data from studies specifically recruiting individuals with NA-resistance at baseline |
| Population | - Clear denominator followed up for a defined period of time<br>- Clear reporting on numbers lost to follow up and reasons<br>- Individual level data on age, sex, HBeAg status, HIV/HCV/HDV coinfection, prior antiviral exposure (duration, agent) |
| Intervention | - Individual level data on: dose, duration, adherence/ gaps in treatment |
| Outcome | - Clear reporting of clinical resistance phenotypes, number attempted to sequence, number successfully sequenced, and number resistant within each group (Supplementary figure 2)<br>- Clear description of which individual/combinations of RAMs were considered significant (and methods used to investigate newly identified polymorphisms)<br>- Description of RAMs as haplotypes, as often combinations confer resistance<br>- Publication of metadata alongside sequence data in public archives |

## Data Availability

All data are available in the supplementary materials

