## Supplementary materials for "A systematic review and meta-analysis of the risk of hepatitis B virus (HBV) genotypic resistance in people treated with entecavir or tenofovir"

### CONTENTS:


**Supplementary tables**

- Supplementary table 1: PRISMA checklist
- Supplementary table 2: Resistance criteria for tenofovir papers
- Supplementary table 3: Characteristics of studies included in the systematic review
- Supplementary table 4: Sensitivity analyses

**Supplementary figures**

- Supplementary figure 1: Summary of risk of bias scores for included studies
- Supplementary figure 2: Data flow to determine risk of clinical and genotypic resistance
- Supplementary figure 3: Relationship between clinical and methodological sources of heterogeneity
- Supplementary figure 4: Funnel plots to investigate publication bias

**Supplementary text**

- Supplementary text 1: Methods - Search strategies
- Supplementary text 2: Methods - Screening and data extraction
- Supplementary text 3: Results: Sensitivity and outlier analysis

**Supplementary references**

### Supplementary table 1: PRISMA checklist[1]

| Section and Topic | Item # | Checklist item | Location where item is reported |
|---|---|---|---|
| **TITLE** | | | |
| Title | 1 | Identify the report as a systematic review. | Title |
| **ABSTRACT** | | | |
| Abstract | 2 | See the PRISMA 2020 for Abstracts checklist. | Abstract |
| **INTRODUCTION** | | | |
| Rationale | 3 | Describe the rationale for the review in the context of existing knowledge. | Intro |
| Objectives | 4 | Provide an explicit statement of the objective(s) or question(s) the review addresses. | Intro |
| **METHODS** | | | |
| Eligibility criteria | 5 | Specify the inclusion and exclusion criteria for the review and how studies were grouped for the syntheses. | Methods |
| Information sources | 6 | Specify all databases, registers, websites, organisations, reference lists and other sources searched or consulted to identify studies. Specify the date when each source was last searched or consulted. | Supplement |
| Search strategy | 7 | Present the full search strategies for all databases, registers and websites, including any filters and limits used. | Supplement |
| Selection process | 8 | Specify the methods used to decide whether a study met the inclusion criteria of the review, including how many reviewers screened each record and each report retrieved, whether they worked independently, and if applicable, details of automation tools used in the process. | Supplement |
| Data collection process | 9 | Specify the methods used to collect data from reports, including how many reviewers collected data from each report, whether they worked independently, any processes for obtaining or confirming data from study investigators, and if applicable, details of automation tools used in the process. | Methods/Supplement |
| Data items | 10a | List and define all outcomes for which data were sought. Specify whether all results that were compatible with each outcome domain in each study were sought (e.g. for all measures, time points, analyses), and if not, the methods used to decide which results to collect. | Methods/Supplement |
| | 10b | List and define all other variables for which data were sought (e.g. participant and intervention characteristics, funding sources). Describe any assumptions made about any missing or unclear information. | Supplement |
| Study risk of bias assessment | 11 | Specify the methods used to assess risk of bias in the included studies, including details of the tool(s) used, how many reviewers assessed each study and whether they worked independently, and if applicable, details of automation tools used in the process. | Methods/Supplement |
| Effect measures | 12 | Specify for each outcome the effect measure(s) (e.g. risk ratio, mean difference) used in the synthesis or presentation of results. | Methods |
| Synthesis methods | 13a | Describe the processes used to decide which studies were eligible for each synthesis (e.g. tabulating the study intervention characteristics and comparing against the planned groups for each synthesis (item #5)). | Methods |
| | 13b | Describe any methods required to prepare the data for presentation or synthesis, such as handling of missing summary statistics, or data conversions. | Methods |
| | 13c | Describe any methods used to tabulate or visually display results of individual studies and syntheses. | Methods |
| | 13d | Describe any methods used to synthesize results and provide a rationale for the choice(s). If meta-analysis was performed, describe the model(s), method(s) to identify the presence and extent of statistical heterogeneity, and software package(s) used. | Methods |
| | 13e | Describe any methods used to explore possible causes of heterogeneity among study results (e.g. subgroup analysis, meta-regression). | Methods |
| | 13f | Describe any sensitivity analyses conducted to assess robustness of the synthesized results. | Methods |
| Reporting bias assessment | 14 | Describe any methods used to assess risk of bias due to missing results in a synthesis (arising from reporting biases). | Methods |
| Certainty assessment | 15 | Describe any methods used to assess certainty (or confidence) in the body of evidence for an outcome. | Methods |

| RESULTS | | | |
|---|---|---|---|
| Study selection | 16a | Describe the results of the search and selection process, from the number of records identified in the search to the number of studies included in the review, ideally using a flow diagram. | Figure 1 |
| | 16b | Cite studies that might appear to meet the inclusion criteria, but which were excluded, and explain why they were excluded. | Discussion |
| Study characteristics | 17 | Cite each included study and present its characteristics. | Table 1 |
| Risk of bias in studies | 18 | Present assessments of risk of bias for each included study. | Table 1 |
| Results of individual studies | 19 | For all outcomes, present, for each study: (a) summary statistics for each group (where appropriate) and (b) an effect estimate and its precision (e.g. confidence/credible interval), ideally using structured tables or plots. | Results figures |
| Results of syntheses | 20a | For each synthesis, briefly summarise the characteristics and risk of bias among contributing studies. | Results |
| | 20b | Present results of all statistical syntheses conducted. If meta-analysis was done, present for each the summary estimate and its precision (e.g. confidence/credible interval) and measures of statistical heterogeneity. If comparing groups, describe the direction of the effect. | Results |
| | 20c | Present results of all investigations of possible causes of heterogeneity among study results. | Results and Supplement |
| | 20d | Present results of all sensitivity analyses conducted to assess the robustness of the synthesized results. | Results and Supplement |
| Reporting biases | 21 | Present assessments of risk of bias due to missing results (arising from reporting biases) for each synthesis assessed. | Results |
| Certainty of evidence | 22 | Present assessments of certainty (or confidence) in the body of evidence for each outcome assessed. | Results |
| **DISCUSSION** | | | |
| Discussion | 23a | Provide a general interpretation of the results in the context of other evidence. | Discussion |
| | 23b | Discuss any limitations of the evidence included in the review. | Discussion |
| | 23c | Discuss any limitations of the review processes used. | Discussion |
| | 23d | Discuss implications of the results for practice, policy, and future research. | Discussion |
| **OTHER INFORMATION** | | | |
| Registration and protocol | 24a | Provide registration information for the review, including register name and registration number, or state that the review was not registered. | Methods |
| | 24b | Indicate where the review protocol can be accessed, or state that a protocol was not prepared. | Methods |
| | 24c | Describe and explain any amendments to information provided at registration or in the protocol. | Methods |
| Support | 25 | Describe sources of financial or non-financial support for the review, and the role of the funders or sponsors in the review. | Funding |
| Competing interests | 26 | Declare any competing interests of review authors. | Competing interests |
| Availability of data, code and other materials | 27 | Report which of the following are publicly available and where they can be found: template data collection forms; data extracted from included studies; data used for all analyses; analytic code; any other materials used in the review. | Data via supplementary file |

### Supplementary table 2: Resistance criteria for TFV papers

| Study ID | 'Established' TFV RAMs in EASL guideline (A181T/V and N236T) (Table 1A) | | 'Putative' RAM combinations (Table 1B) | |
|---|---|---|---|---|
| | Identified | Classified as resistant | Identified | Classified as resistant |
| **Both established and/or putative RAMs classified as resistant** | | | | |
| Bakhshizadeh 2015 | N | Y | N | Y (A194T) |
| Hongthanakorn 2011 | N | Y | N | Y (A194T) |
| Patterson 2011 | Y | Y | NS | NS |
| Tan 2008 | Y | Y | NS | NS |
| Liang 2019 | N | Y | L180M+M204I/V +V173L | N |
| DeFrancesco 2015 | N | N | 1) A194T, 2)L180M+M204I/V +V173L | 1) Y (A194T), 2) N |
| **Neither established nor putative RAMs classified as resistant** | | | | |
| Corsa 2014 | Y | N | A194A/P | N |
| Hou 2015 | Y | N | (V173L/V/L180M/L/M204V/M/S213T) | N |
| Boyd 2014 | N | N (Geno2pheno*) | L180M+M204I/V +V173L | N (Geno2pheno*) |
| Snow-Lampart 2011 | N | N | L180M+M204V +V173L | N |
| Stephan 2005 | NS | N | L180M+M204V +V173L | N |
| Boyd 2015 | N | NS | L180M+M204I/V +V173L | N |
| Singla 2015 | N | N (Geno2pheno*) | N | N (Geno2pheno*) |
| **Established RAMs not classified as resistant, putative not reported** | | | | |
| Cathcart 2018 | Y | N | N | NS |
| Lim 2016 (2) | Y (all A181V/T and/or N236T at baseline) | N | N | NS |
| Yang 2015 | Y at baseline | N | NS | NS |
| Berg 2014 | Y | N | NS | NS |
| Liu 2017 | Y | N | NS | NS |
| **RAM criteria not specified, RAM details not reported** | | | | |
| Lim 2016 (1) | Y (individuals excluded if identified at baseline) | NS | N | NS |
| Fung 2017 | N | NS | N | NS |
| Marcellin 2008 | NS | NS | NS | NS |
| Pan 2014 | NS | NS | NS | NS |
| Chan 2023 | NS | NS | NS | NS |
| Srivastava 2016 | NS | NS | NS | NS |
| Buti 2015 | NS | NS | NS | NS |
| Cho 2015 | NS | NS | NS | NS |

*NB HBV geno2pheno does not list any TFV RAMs

**Supplementary table 3: Characteristics of 62 studies included in the systematic review.** Where multiple subgroups of individuals are reported in the study, only subgroups meeting inclusion criteria are detailed in the table.

Abbreviations: ART = antiretroviral therapy, eAg- = HBeAg negative, eAg+ = HBeAg positive, EE = NA experienced on Entecavir, ET = NA experienced on tenofovir, GR = genotypic resistance, NA = nucleoside analogue, NE = NA naive on Entecavir, NT = NA naive on tenofovir, OD = once a day, Prosp = prospective, Retro = retrospective, TAF = tenofovir alafenamide, TDF = tenofovir disoproxil. VBT = virological breakthrough, * RAM identified for any NA

| Study | | | | Intervention | | Population | | | | | Outcome | | Bias |
|---|---|---|---|---|---|---|---|---|---|---|---|---|---|
| **Study** | **Study design** | **WHO region** | **Study size and group** | **Drug/ Dose** | **Duration (m)** | **Prior NA exposure** | **Baseline GR*** | **Age group** | **HBeAg status** | **HIV co-infection** | **Seq. criteria** | **GR risk (%)** | **Risk of bias (1 = high, 5 = low)** |
| Bakhshizadeh 2015[2] | Cohort/cross sectional; Prosp. | EMR | 93 NT | TDF 300mg OD | mean 21.10 (R 6-36) | Naïve | NA | Adults | eAg+ and eAg- | No | All | 0.0 | 5 |
| Berg 2014[3] | Clinical trial; Prospective | AMR, EMR | 53 ET | TDF 300mg OD | EP 38.00 | Experienced | All | Adults | eAg+ and eAg- | Not reported | VBT only | 0.0 | 2 |
| Boyd 2014[4] | Cohort/cross sectional; Prosp. | EUR | 24 ET | TDF (as ART) | med 74.70 (IQR 33.4-94.7) | Experienced | Not reported | Adults | eAg+ and eAg- | Yes | Unclear | 0.0 | 4 |
| Boyd 2015[5] | Cohort/cross sectional; Prosp. | AFR | 86 NT | TDF (as ART) | med 24.30 (IQR 17.6-30.8) | Naïve | NA | Adults | eAg+ and eAg- | Yes | All | 0.0 | 5 |
| Buti 2015[6] | Clinical trial; Prospective | AMR, EUR, WPR | 585 NT | TDF 300mg OD | EP 77.3 | Naïve | NA | Adults | eAg+ and eAg- | Not reported | All | 0.0 | 3 |
| Cathcart 2018[7] | Clinical trial; Prospective | AMR, SEAR, EUR, WPR | 1002 NT 296 ET | TAF 25mg, TDF 300mg | EP 22.09 | Naïve and experienced | Mix | Adults | eAg+ and eAg- | No | All | 0.0 NT 0.0 ET | 5 |
| Chan 2023[8] | Clinical trial; Prospective | AMR, SEAR, EUR, EMR, WPR | 993 NT 305 ET | TAF 25mg, TDF 300mg | EP 55.2 | Naïve and experienced | Not reported | Adults | eAg+ and eAg- | No | All | 0.0 NT 0.0 ET | 4 |
| Chang 2005[9] | Clinical trial; Prospective | AMR, EUR, EMR, WPR | 39 EE | ETV 1mg OD | EP 11.04 | Experienced | All | Adults | eAg+ and eAg- | No | All | 0.0 | 4 |
| Chang 2006[10] | Clinical trial; Prospective | AMR, SEAR, EUR, WPR | 344 NE 10 EE | ETV 0.5 mg OD | EP 11.96 | Naïve and experienced | NA | Adults | eAg+ and eAg- | No | All | 0.0 NE 0.0 EE | 4 |
| Chang 2009[11] | Clinical trial; Prospective | AMR, SEAR, EUR, WPR | 243 NE | ETV 0.5 mg OD | EP 24.00 | Naïve | NA | Adults | all eAg+ | No | All | 0.0 | 4 |
| Chen 2011[12] | Cohort/cross sectional; Prosp. | WPR | 48 NE | ETV 0.5 mg OD | EP 24.00 | Naïve | NA | Adults | eAg+ and eAg- | No | VBT only | 0.0 | 5 |
| Cho 2015[13] | Cohort or cross sectional; Retros | WPR | 146 EE 56 ET | ETV 0.5/1mg OD. TDF 300mg OD | med 37.70 (IQR 23.4-74.5) | Experienced | No | Adults | eAg+ and eAg- | No | All | 8.9 EE 0.0 ET | 4 |
| Cho 2017[14] | Cohort/cross sectional; Prosp. | WPR | 1009 NE | ETV 0.5 mg OD | med 26.50 (R 6-77.4) | Naïve | NA | Adults | eAg+ and eAg- | No | Unclear | 1.2 | 4 |
| Corsa 2014[15] | Clinical trial; Prospective | AMR, EUR, WPR | 280 ET | TDF 300mg OD | EP 22.09 | Experienced | All | Adults | eAg+ and eAg- | No | All | 0.0 | 5 |
| DeFrancesco 2015[16] | Cohort or cross sectional; Retrospective | EUR | 11 ET | TDF 300mg OD | EP 24.00 | Experienced | All | Adults | eAg+ and eAg- | Not reported | VBT only | 9.1 | 3 |
| Deng 2013[17] | Cohort/cross sectional; Prosp. | WPR | 21 EE | ETV 1mg OD | EP 33.14 | Experienced | All | Adults | all eAg+ | Not reported | All | 23.4 | 3 |
| Fung 2014[18] | Clinical trial; Prospective | AMR, EUR, WPR | 141 ET | TDF 300mg OD | EP 22.09 | Experienced | All | Adults | eAg+ and eAg- | No | All | 0.0 | 5 |
| Fung 2017[19] | Clinical trial; Prospective | AMR, EUR, WPR | 141 ET | TDF 300mg OD | EP 55.23 | Experienced | All | Adults | eAg+ and eAg- | No | All | 0.0 | 5 |

| | | | | | | | | | | | | | |
|---|---|---|---|---|---|---|---|---|---|---|---|---|---|
| Gish 2007[20] | Clinical trial; Prospective | AMR | 243 NE | ETV 0.5 mg OD | EP 22.09 | Naïve | NA | Adults | all eAg+ | Not reported | All | 0.0 | 5 |
| Gwak 2013[21] | Cohort or cross sectional; Prospective | WPR | 58 NE | ETV 0.5 mg OD | EP 24.00 | Naïve | NA | Adults | eAg+ and eAg- | No | VBT only | 0.0 | 5 |
| Ha 2011[22] | Cohort or cross sectional; Retros | AMR | 107 NE | ETV 0.5 mg OD | EP 48.00 | Naïve | NA | Adults | all eAg- | No | VBT only | 0.0 | 5 |
| Heathcote 2011[23] | Clinical trial; Prospective | AMR, EUR, WPR | 196 ET | TDF 300mg OD | EP 22.09 | Experienced | No | Adults | eAg+ and eAg- | No | All | 0.0 | 5 |
| Heo 2012[24] | Clinical trial; Prospective | WPR | 34 EE | ETV 1mg OD | EP 22.09 | Experienced | No | Adults | all eAg+ | No | All | 2.9 | 5 |
| Hongthanakorn 2011[25] | Cohort or cross sectional; Retros | AMR | 15 ET<br>43 NE<br>13 EE | ETV, TDF, dose NS | mean 37.50 ET<br>37.50 NE<br>37.5 EE<br>(R 12-102) | Naïve and experienced | Not reported | Adults | eAg+ and eAg- | No | VBT only | 0.0 ET<br>2.3 NE<br>38.5 EE | 4 |
| Hou 2015[26] | Clinical trial; Prospective | WPR | 244 NT<br>11 ET | TDF 300mg OD | EP 11.04 | Naïve and experienced | Mix | Adults | eAg+ and eAg- | No | All | 0.4 NT<br>0.0 ET | 4 |
| Jonas 2016[27] | Clinical trial; Prospective | AMR, SEAR, EUR | 120 NE | ETV 0.015 mg/kg OD, max 0.5 mg | 22.09 | Naïve | NA | Children | all eAg+ | No | All | 3.3 | 4 |
| Kamezaki 2011[28] | Cohort or cross sectional; Retro | WPR | 81 NE | ETV 0.5 mg OD | mean 27.00 | Naïve | NA | Adults | eAg+ and eAg- | No | VBT only | 2.5 | 5 |
| Kamezaki 2013[29] | Cohort or cross sectional; Retro | WPR | 135 NE | ETV 0.5 mg OD | 26.90<br>(SD +/- 321.6) | Naïve | NA | Adults | eAg+ and eAg- | No | Unclear | 1.5 | 5 |
| Karino 2010[30] | Cohort or cross sectional; Prosp | WPR | 42 EE | ETV 1mg OD | EP 34.06 | Experienced | All | Adults | eAg+ and eAg- | No | All | 30.9 | 3 |
| Kim 2010 (1)[31] | Cohort or cross sectional; Retro | WPR | 73 NE | ETV 0.5 mg OD | mean 18.40 (SD +/- 3.8) | Naïve | NA | Adults | eAg+ and eAg- | No | VBT only | 0.0 | 4 |
| Kim 2010 (2)[32] | Cohort or cross sectional; Retro | WPR | 24 EE | ETV 1mg OD | EP 24.00 | Experienced | All | Adults | eAg+ and eAg- | No | VBT only | 25.0 | 5 |
| Kim 2017[33] | Cohort or cross sectional; Retro | WPR | 202 NE<br>56 EE | ETV 0.5 mg OD | median 59 NE/ EE (R9-101) | Naïve and experienced | No | Adults | eAg+ and eAg- | No | VBT only | 3.0 NE<br>3.6 EE | 4 |
| Kitrinos 2014[34] | Clinical trial; Prospective | AMR, EUR, WPR | 389 ET | TDF 300mg OD | EP 72.00 | Experienced | Mix | Adults | eAg+ and eAg- | No | All | 0.0 | 3 |
| Liang 2019[35] | Clinical trial; Prospective | WPR | 257 NT | TDF 300mg OD | EP 55.00 | Naive | NA | Adults | eAg+ and eAg- | No | All | 0.0 | 5 |
| Lim 2016 (1)[36] | Clinical trial; Prospective | WPR | 44 ET | TDF 300mg OD | EP 11.09 | Experienced | All | Adults | eAg+ and eAg- | No | All | 0.0 | 5 |
| Lim 2016 (2)[37] | Clinical trial; Prospective | WPR | 50 ET | TDF 300mg OD | EP 22.09 | Experienced | All | Adults | eAg+ and eAg- | No | All | 0.0 | 5 |
| Liu 2016[38] | Cohort/cross sectional; Prosp. | WPR | 33 NE<br>56 EE | ETV 0.5mg naive, 1mg experienced | 69.00 NE (R 60-75), 57.00 EE (R12-75) | Naïve and experienced | Mix | Adults | eAg+ and eAg- | No | VBT only | 0.0 NE<br>16.1 EE | 5 |
| Liu 2017[39] | Clinical trial; Prospective | AMR, EUR, WPR | 426 ET | TDF 300mg OD | EP 96.00 | Experienced | Mix | Adults | eAg+ and eAg- | No | All | 0.0 | 4 |
| Lok 2012[40] | Clinical trial; Prospective | ARF, AMR, EUR, SEAR, WPR | 182NE | ETV 0.5 mg OD | EP 23.01 | Naïve | NA | Adults; Children | eAg+ and eAg- | Not reported | All | 0.0 | 3 |
| Marcellin 2008[41] | Clinical trial; Prospective | AMR, EUR, WPR | 375 NT<br>51 ET | TDF 300mg OD | EP 11.04 | Naïve and experienced | Not reported | Adults | eAg+ and eAg- | No | All | 0.0 NT<br>0.0 ET | 4 |
| Pan 2014[42] | Clinical trial; Prospective | AMR | 87 NT | TDF 300mg OD | EP 11.04 | Naïve | NA | Adults | eAg+ and eAg- | Not reported | All | 0.0 | 5 |
| Park 2011[43] | Cohort or cross sectional; Prospective | WPR | 55 EE | ETV 1mg OD | median 24.0 (12-47) | Experienced | All | Adults | eAg+ and eAg- | No | VBT only | 36.4 | 5 |
| Patterson 2011[44] | Clinical trial; Prospective | WPR | 38 ET | TDF 300mg OD | EP 22.09 | Experienced | All | Adults | eAg+ and eAg- | No | VBT only | 7.9 | 5 |
| Sherman 2006[45] | Clinical trial; Prospective | AMR, SEAR, EUR, EMR, WPR | 141 ET | ETV 1mg OD | EP 11.05 | Experienced | All | Adults | all eAg+ | No | All | 1.4 | 4 |

| | | | | | | | | | | | | | |
|---|---|---|---|---|---|---|---|---|---|---|---|---|---|
| Shin 2011[46] | Cohort or cross sectional; Retro | WPR | 61 NE | ETV 0.5mg OD | EP 11.05 | Naïve | NA | Adults | eAg+ and eAg- | No | VBT only | 0.0 NE | 4 |
| Singla 2015[47] | Cohort/cross sectional; Prosp. | SEAR | 30 NT<br>39 NE | TDF 300mg OD<br>ETV 0.5mg OD | EP 12.00 | Naïve | NA | Adults | eAg+ and eAg- | Not reported | All | 0.0 NT<br>0.0 NE | 3 |
| Snow-Lampart 2011[48] | Clinical trial; Prospective | AMR, EUR, WPR | 196 TE | TDF 300mg OD | EP 22.09 | Experienced | Mix | Adults | eAg+ and eAg- | No | All | 0.0 | 4 |
| Srivastava 2016[49] | Cohort/cross sectional; Prosp. | SEAR | 26 NT<br>25 NE | ETV 0.5mg OD<br>TDF 300mg OD | EP 24.00 | Naïve | NA | Adults; Children | eAg+ and eAg- | No | All | 0.0 NT<br>0.0 NE | 4 |
| Stephan 2005[50] | Cohort or cross sectional; Retro | EUR | 24 ET | TDF (as ART) | EP 11.05 | Experienced | Not reported | Adults | eAg+ and eAg- | Yes | All | 0.0 | 5 |
| Suzuki 2008[51] | Clinical trial; Prospective | WPR | 84 EE | ETV 0.5mg, 1mg OD | EP 12.00 | Experienced | All | Adults | eAg+ and eAg- | No | VBT only | 0.0 | 3 |
| Suzuki 2010[52] | Cohort or cross sectional; Retro | WPR | 130 EE | ETV 0.5 mg OD | EP 12.00 | Experienced | All | Adults | eAg+ and eAg- | Not reported | Unclear | 0.8 | 5 |
| Suzuki 2019[53] | Cohort or cross sectional; Retro | WPR | 1094 NE | ETV 0.5mg | median 66.00 (R 12 - 120) | Naïve | NA | Adults | eAg+ and eAg- | No | VBT only | 0.7 | 5 |
| Tan 2008[54] | Cohort or cross sectional; Retro | AMR | 10 ET | TDF 300mg OD | EP 20.50 | Experienced | Mix | Adults | eAg+ and eAg- | Not reported | All | 10.0 | 3 |
| Tenney 2007[55] | Cohort/cross sectional; Prosp. | AMR, EUR, WPR | 192 EE | ETV 1mg OD | EP 24.00 | Experienced | All | Adults | eAg+ and eAg- | Not reported | All | 12.0 | 2 |
| Tenney 2009[56] | Clinical trial; Prospective | AMR, SEAR, EUR, WPR | 108 NE<br>33 EE | ETV 0.5mg naive, 1mg exp | EP 60.00 | Naïve and experienced | All | Adults | eAg+ and eAg- | Not reported | All | 8.3 NE<br>42.4 EE | 4 |
| Tsai 2012[57] | Cohort/cross sectional; Prosp. | WPR | 98 NE | ETV 0.5 mg OD | EP 11.05 | Naïve | NA | Adults | eAg+ and eAg- | Not reported | VBT only | 0.0 NE | 4 |
| Wang 2013[58] | Cohort/cross sectional; Prosp. | WPR | 25 EE | ETV 0.5 mg OD | EP 12.00 | Experienced | No | Adults | all eAg+ | No | All | 4.0 | 3 |
| Xu 2022[59] | Clinical trial; Prospective | WPR | 208 NE<br>18 EE | ETV 0.5mg OD | EP 44.00 | Naïve and experienced | Mix | Adults | eAg+ and eAg- | Not reported | All | 1.4 NE<br>22.2 EE | 5 |
| Yang 2015[60] | Cohort or cross sectional; Retro | WPR | 28 ET | TDF 300mg OD | EP 11.05 | Experienced | All | Adults | eAg+ and eAg- | No | All | 0.0 ET | 5 |
| Yokosuka 2010[61] | Clinical trial; Prospective | WPR | 66 NE | ETV 0.01, 0.1, 0.5mg then 0.5mg rollover | EP 27.6 | Naïve | NA | Adults | eAg+ and eAg- | No | All | 1.5 | 5 |
| Yuen 2015[62] | Clinical trial; Prospective | WPR | 30 NE | ETV 0.5 mg OD | EP 22.09 | Naïve | NA | Adults | eAg+ and eAg- | No | All | 0.0 | 5 |
| Zhou 2017[63] | Cohort/cross sectional; Prosp. | WPR | 33 ET | TDF 300mg OD | EP 11.05 | Experienced | All | Adults | eAg+ and eAg- | No | Unclear | 0.0 ET | 5 |

**Supplementary table 4A: Sensitivity analysis for Experienced/Tenofovir (n=19)**

| Year group* | Number of studies remaining | Pooled risk estimate from random effects model* | 95% CI lower | 95% CI upper | Heterogeneity ($I^2$) |
|---|---|---|---|---|---|
| Baseline analysis | | | | | |
| 2 | 8 | 0.000 | 0.000 | 0.006 | 66% |
| Study size >30 | | | | | |
| 2 | 6 | 0.000 | 0.000 | 0.005 | 56% |
| Exclude studies with high risk of bias | | | | | |
| 2 | 8 | 0.000 | 0.000 | 0.006 | 66% |
| Study design = clinical trial | | | | | |
| 2 | 6 | 0.000 | 0.000 | 0.005 | 56% |
| Sequencing criteria = All sequenced | | | | | |
| 2 | 6 | 0.000 | 0.000 | 0.000 | 4% |
| Baseline mutations - exclude those where all individuals have RAMs at baseline | | | | | |
| 2 | 4 | 0.000 | 0.000 | 0.000 | 39% |

* Treatment duration (years)

** Or single study estimate if only 1 study remaining for analysis

**Supplementary table 4B: Sensitivity analysis for Naive/Entecavir (n = 22)**

| Year group | Number of studies remaining | Pooled risk estimate from random effects model* | 95% CI lower | 95% CI upper | Heterogeneity ($I^2$) |
|---|---|---|---|---|---|
| Baseline analysis | | | | | |
| 2 | 13 | 0.003 | 0.000 | 0.009 | 41% |
| 4 | 2 | 0.006 | 0.000 | 0.027 | 47% |
| ≥5 | 4 | 0.009 | 0.001 | 0.023 | 49% |
| Study size >30 | | | | | |
| 2 | 11 | 0.004 | 0.000 | 0.011 | 51% |
| 4 | 2 | 0.006 | 0.000 | 0.027 | 47% |
| ≥5 | 4 | 0.009 | 0.001 | 0.023 | 49% |
| Study design = clinical trial | | | | | |
| 2 | 6 | 0.002 | 0.000 | 0.013 | 60% |
| 4 | 1 | 0.014 | 0.003 | 0.042 | NA |
| ≥5 | 1 | 0.009 | 0.000 | 0.051 | NA |
| Sequencing criteria = All sequenced | | | | | |
| 2 | 7 | 0.001 | 0.000 | 0.010 | 53% |
| 4 | 1 | 0.014 | 0.003 | 0.042 | NA |
| ≥5 | 1 | 0.009 | 0.000 | 0.051 | NA |
| Baseline mutations - exclude those where all individuals have RAMs at baseline | | | | | |
| 2 | 13 | 0.003 | 0.000 | 0.009 | 41% |
| 4 | 2 | 0.006 | 0.000 | 0.027 | 47% |
| ≥5 | 3 | 0.009 | 0.000 | 0.031 | 65% |

* Treatment duration (years)

** Or single study estimate if only 1 study remaining for analysis

#### Supplementary table 4C: Sensitivity analysis for Experienced/Entecavir (n = 18)

| Year group | Number of studies remaining | Pooled risk estimate from random effects model* | 95% CI lower | 95% CI upper | Heterogeneity ($I^2$) |
|---|---|---|---|---|---|
| Baseline analysis | | | | | |
| 2 | 4 | 0.170 | 0.054 | 0.329 | 87% |
| 3 | 4 | 0.226 | 0.082 | 0.410 | 82% |
| ≥5 | 2 | 0.201 | 0.016 | 0.501 | 93% |
| Study size >30 | | | | | |
| 2 | 3 | 0.151 | 0.026 | 0.342 | 90% |
| 3 | 2 | 0.180 | 0.020 | 0.439 | 91% |
| ≥5 | 3 | 0.201 | 0.016 | 0.501 | 93% |
| Exclude studies with high risk of bias | | | | | |
| 2 | 3 | 0.193 | 0.023 | 0.454 | 88% |
| 3 | 4 | 0.226 | 0.082 | 0.410 | 82% |
| ≥5 | 3 | 0.201 | 0.016 | 0.501 | 93% |
| Sequencing criteria = All sequenced | | | | | |
| 2 | 2 | 0.181 | 0.017 | 0.179 | 62% |
| 3 | 3 | 0.193 | 0.064 | 0.476 | 84% |
| ≥5 | 1 | 0.515 | 0.335 | 0.692 | NA |
| Baseline mutations - exclude those where all individuals have RAMs at baseline | | | | | |
| 2 | 1 | 0.029 | 0.001 | 0.153 | NA |
| 3 | 2 | 0.193 | 0.000 | 0.545 | 85% |
| ≥5 | 2 | 0.089 | 0.005 | 0.244 | 80% |

* Treatment duration (years)

** Or single study estimate if only 1 study remaining for analysis

**Supplementary figure 1: Summary of risk of bias scores for included studies.**

Percentage of studies rated as “yes” (green), “unclear” (orange) or “no” (red) to five quality Joanna Briggs Institute bias scoring questions:

Q1. Were the criteria for inclusion in the study clearly defined?
Q2. Were the study subjects and the setting described in detail?
Q3. Was the exposure (drug treatment) measured in a valid and reliable way?
Q4. Were objective, standard criteria used for measurement of the condition (ie. chronic HBV)?
Q5. Were the outcomes measured in a valid and reliable way?

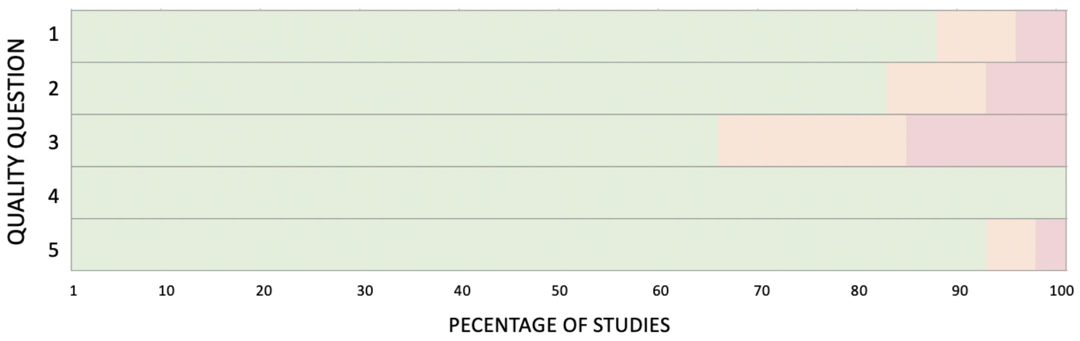

**Supplementary figure 2: Gold standard data flow to determine risk of clinical and genotypic resistance.**

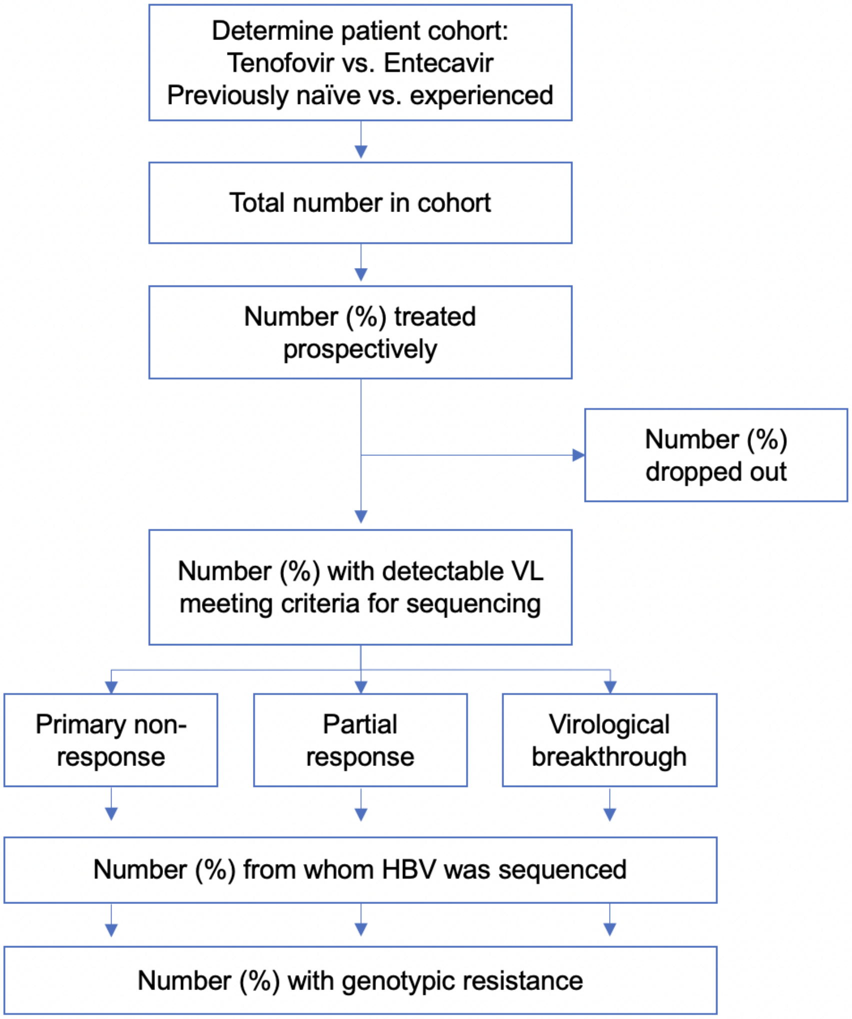

**Supplementary figure 3: Relationship between clinical and methodological sources of heterogeneity (publication date, HBeAg status, HIV status, study type, publication date, exclusion of those with adherence issues, baseline resistance status and sequencing criteria) and the observed risk of genotypic resistance over time.** All 62 studies are shown without deduplication of cohorts where the same data is published multiple times.

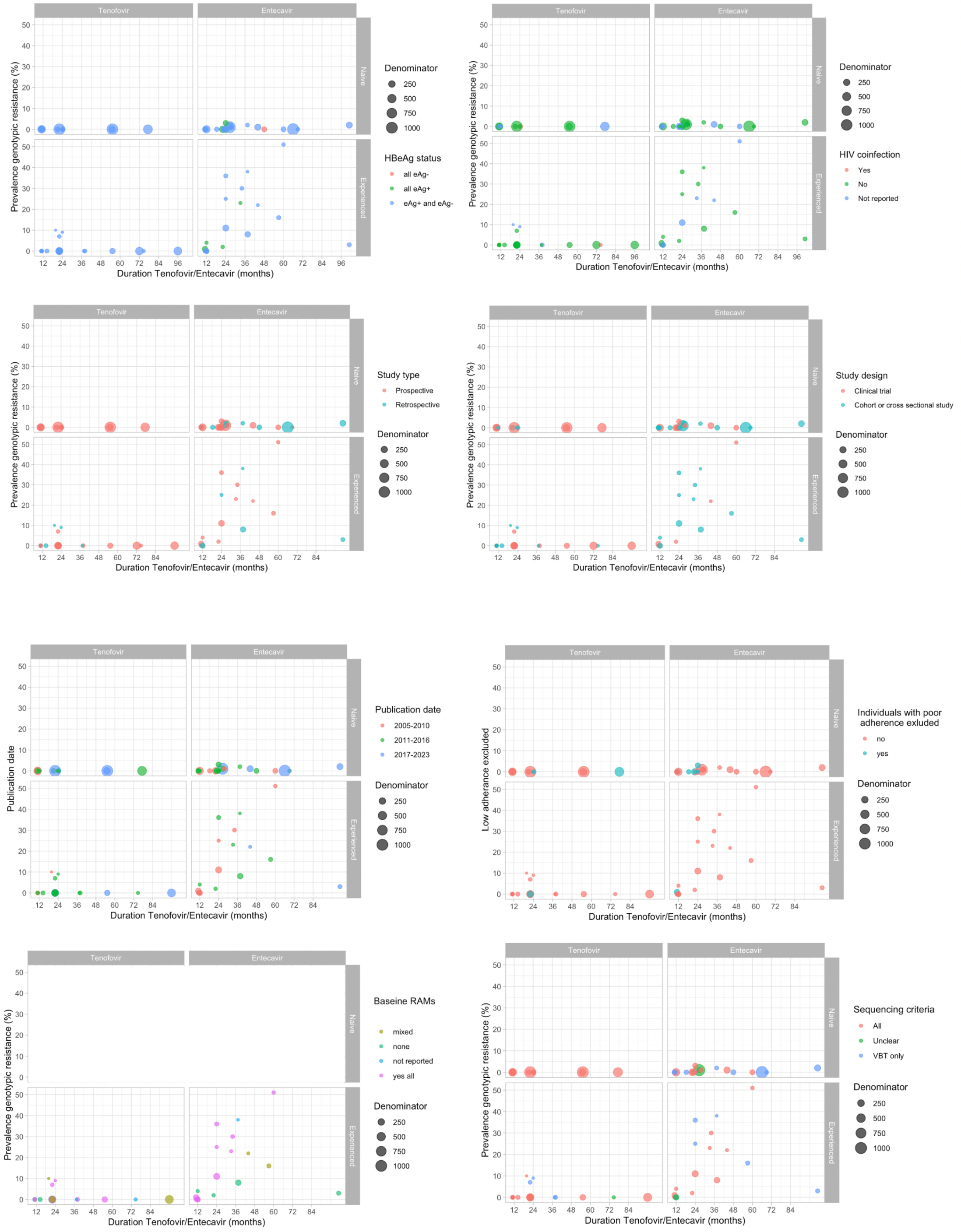

**Supplementary figure 4: Funnel plots to investigate publication bias**

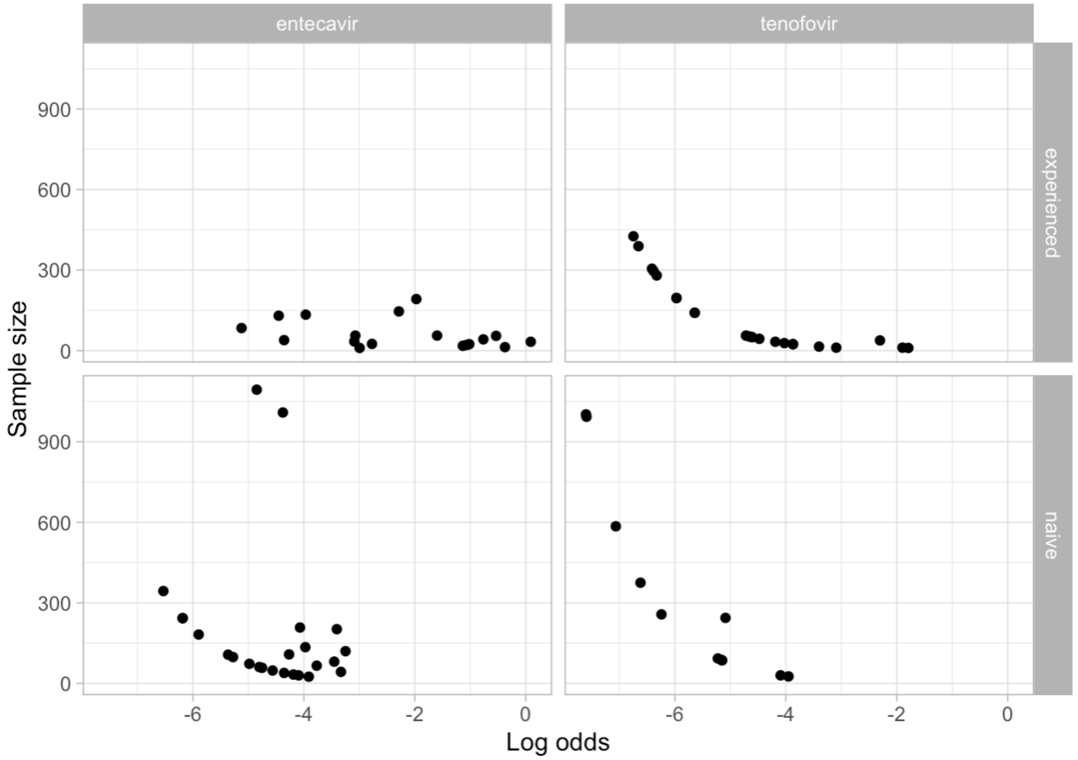

**Supplementary text 1:**

**Methods - Search strategies**

**PubMed**

((((("Hepatitis B virus"[Mesh]) OR ("hepatitis b"[Title/Abstract] OR CHB[Title/Abstract] OR HPV[Title/Abstract])) AND ((("Entecavir" [Supplementary Concept]) OR "Tenofovir"[Mesh]) OR (Entecavir[Title/Abstract] OR Tenofovir[Title/Abstract] OR Viread[Title/Abstract])))) AND (("Drug Resistance"[Mesh]) OR (resistan*[Title/Abstract] OR "drug mutat*"[Title/Abstract] OR DRMs[Title/Abstract] OR escap*[Title/Abstract] OR mutat*[Title/Abstract]))) AND (chronic*)

*Filters applied: English.*

---

**Database: Embase 1974 to present**

Link to search history: https://ovidsp.ovid.com/ovidweb.cgi?T=JS&NEWS=N&PAGE=main&SHAREDSEARCHID=4n05jJsL1ji4FTF87qkNYBsZHPU9nRUTS2dvJ0VY0eGxMFI8LbocaxjpUQAhZZhHX

**Search Strategy:**

**1** exp Hepatitis B virus/ (62830)

**2** ("hepatitis b" or CHB or HBV).ti,ab. (149344)

**3** 1 or 2 (158457)

**4** Entecavir/ (11052)

**5** Tenofovir/ (21517)

**6** (Entecavir or Tenofovir or Viread).ti,ab. (20044)

**7** 4 or 5 or 6 (35130)

**8** exp drug resistance/ (374437)

**9** (resistan* or "drug mutat*" or DRMs or escap* or mutat*).ti,ab. (2570076)

**10** 8 or 9 (2642215)

**11** exp chronic viral hepatitis/ (32355)

**12** chronic*.ti,ab,kw. (2067571)

**13** 11 or 12 (2072437)

**14** 3 and 7 and 10 and 13 (2870)

**15** limit 14 to english language (2649)

---

**Database: Global Health <1973 to 2023 Week 21>**

Link to search history: https://ovidsp.ovid.com/ovidweb.cgi?T=JS&NEWS=N&PAGE=main&SHAREDSEARCHID=4oiBkIUlltcJfuJ4tgWl6jFpYipaXpXfRQ03vmUTgnLPITNDPCMrv6Kg3xNZrX6Kp

**Search Strategy:**

**1** exp hepatitis b virus/ (39525)

**2** ("hepatitis b" or CHB or HBV).ti,ab. (42432)

**3** 1 or 2 (43954)

**4** Entecavir/ (1174)

**5** Tenofovir/ (3980)

**6** (Entecavir or Tenofovir or Viread).ti,ab. (5825)

**7** 4 or 5 or 6 (6014)

**8** exp drug resistance/ (139291)

**9** (resistan* or "drug mutat*" or DRMs or escap* or mutat*).ti,ab. (347385)

**10** 8 or 9 (355161)

**11** chronic infections/ (23637)

**12** chronic*.ti,ab. (222202)

**13** 11 or 12 (222679)

**14** 3 and 7 and 10 and 13 (615)

**15** limit 14 to english language (539)

---

#### Scopus

( TITLE-ABS-KEY ( "hepatitis b" OR chb OR hbv ) AND TITLE-ABS-KEY ( Entecavir OR Tenofovir OR viread ) AND TITLE-ABS-KEY ( resistan* OR "drug mutat*" OR drms OR escap* OR mutat* ) AND TITLE-ABS-KEY ( chronic* ) ) AND ( LIMIT-TO ( LANGUAGE , "English" ) )

#### Cochrane Central Register of Controlled Trials

#### Issue 6 of 12, June 2023

| | | |
|---|---|---|
| #1 | ("hepatitis b" or CHB or HBV):ti,ab,kw | 10780 |
| #2 | (Entecavir or Tenofovir or Viread):ti,ab,kw | 4649 |
| #3 | (resistan* or "drug mutat*" or DRMs or escap* or mutat*):ti,ab,kw | 103427 |
| #4 | chronic*:ti,ab,kw | 178032 |
| #5 | #1 and #2 and #3 and #4 | 425 |

**Clinicaltrials.gov**

Condition or disease: Hepatitis B, Chronic

Other terms: Entecavir

Condition or disease: Hepatitis B, Chronic

Other terms: Tenofovir

**ISRTCN Registry**

Condition: chronic hepatitis b

Interventions: Entecavir

https://www.isrctn.com/search?q=&filters=condition%3Achronic+hepatitis+b%2Cintervention%3AEntecavir

Condition: chronic hepatitis b

Interventions: Tenofovir

https://www.isrctn.com/search?q=&filters=condition%3Achronic+hepatitis+b%2Cintervention%3ATenofovir

**WHO International Clinical Trials Registry Platform**

https://trialsearch.who.int/AdvSearch.aspx

Title: resistance

Condition: chronic hepatitis b

Intervention: Entecavir

ALL trials

Title: resistance

Condition: chronic hepatitis b

Intervention: Tenofovir

ALL trials

**Supplementary text 2:**
**Methods**

**Screening and data extraction**

Our team first undertook screening of titles and abstracts, then full text review for data extraction, with at least two reviewers independently screening at each stage. In cases where consensus could not be achieved, a third reviewer was asked for consensus. In the case of uncertainty, final decisions on eligibility and extraction were taken by the first and senior author to provide consistency. Reviewers had access to the full paper and were therefore not blind to the author or journal information.

Two review authors independently extracted information for each of the eligible studies after training and piloting the Covidence data extraction tool before use. All individual characteristics (e.g. age, HIV status, HBeAg status) were reported at baseline (ie. on initiation of TFV/ETV). Missing or unclear data recorded as ‘unclear’, ‘missing’ or ‘other’. Data fields collected were:

- Study ID
- Title of paper
- Digital online identifier
- WHO region
- Study design (clinical trial, or cohort/cross-sectional)
- Study type (prospective or retrospective)
- Population description
- Adults or children
- Study setting
- Antiviral regimen including dosage
- Prior antiviral exposure and evidence of phenotypic/genotypic resistance
- Criteria for resistance testing
- Reported incomplete adherence
- HIV coinfection
- HBeAg status at baseline
- Cohort size (denominator on an intention to treat basis)
- Demographics: age, sex
- Duration of treatment (trial endpoint, or mean/median treatment duration for cohorts/cross-sectional studies)

- Sequencing method
- Genotypes reported
- Details of individual RAMs detected
- Prior publication of cohort

The key outcome measure sought was the number of individuals developing genotypic resistance. We also collected data on the number of individuals with clinical resistance and the number successfully sequenced (Supplementary figure 2, supplementary text 2). All included studies looked for genotypic resistance in individuals with VBT. Some studies looked for genotypic resistance in all viraemic individuals (ie. individuals with VBT, primary non-response and partial response). The impact of sequencing criteria on risk estimates was explored in the sensitivity analysis.

Where resistance risk was reported at multiple time points within one paper, the latest time point was collected. If genotypic resistance risk was not reported for naive and experienced individuals separately for the entire trial duration, data from the latest time point at which naive/experienced individuals were reported separately was used. Where multiple doses of NA were compared, data for the cohort receiving the current licensed dose was extracted (typically 300mg/day of TDF, 25mg TAF, 0.5mg ETV (if NA naive) or 1mg ETV (in the setting of LAM resistance). Where outcomes were not described for each dose regimen separately, the pooled results were extracted and regimen noted. Where TAF and TDF were being compared, risk of resistance was pooled. Where study details were not available in the full text or supplementary materials, we reviewed previously published reports of the same study cohort to complete the metadata.

Risk of bias was assessed independently by two reviewers using a modified five question Joanna Briggs Institute quality assessment tool. An answer of “Yes” to all 5 questions equated to a high quality study with low risk of bias. Papers scoring 4 or 3 “Yes” answers ranked as moderate quality/moderate risk of bias and 2 “Yes” as low quality/high risk of bias. Papers scoring only 1 “Yes” were considered very low quality/ high risk of bias and were excluded. Risk of publication bias was alleviated by a comprehensive search strategy.

We analysed our data in four groups determined a priori. In order to decide which studies were eligible for each synthesis, the study population (naive vs. experienced) and NA (ETV vs. TFV) were compared against the planned groups for each synthesis.

**Software**

We exported all references to EndNote 20 (Thomson Reuters, New York, NY) and uploaded to Covidence.org. We removed duplicates automatically through Covidence.org and manually if we identified any further duplicates. We used Covidence software for study selection and data extraction.

**Supplementary text 3:**
**Results: Sensitivity and outlier analysis**

We explored study heterogeneity arising from clinical and methodological diversity by performing sensitivity analyses where heterogeneity in the primary analysis was present (ie. where pooled estimate was generated and $I^2 \neq 0\%$). We also performed a qualitative assessment of outliers.

**i) Naive/Tenofovir**

In NA naive individuals on TFV, heterogeneity ($I^2$) was 0% at all time points, therefore sensitivity analyses were not performed. Only one individual in one study[26] was reported to have genotypic TFV resistance. This individual was reported as nucleos(t)ide naive, but the HBV sequence harboured V173L/V, L180M/L, M204V/M and S213T at baseline, which persisted at week 48 on treatment (putative TFV RAMs Table 2, Supplementary table 3).

**ii) Experienced/Tenofovir**

In NA experienced individuals treated with TFV, heterogeneity was only seen at the 2 year time point ($I^2$ = 66%). Restricting studies to those where all individuals were sequenced led to the largest reduction in heterogeneity to 4%, restricting by study size reduced $I^2$ to 56%, by study design to 56% and by excluding those where all had baseline RAMs to other NAs to 39%, implying multiple factors impacted on heterogeneity (Supplementary table 3A).

One study [44] was classified as an outlier, with its 95% CI not overlapping with the CI of the pooled estimate. It was a small study performed in Australia 2006-2008, 3/38 individuals developed evidence of genotypic resistance, two individuals had N236T mutation at baseline which persisted on TDF treatment, one individual developed A181T and N236T on treatment (accepted TFV RAMs (Table 1A), Supplementary table 2A)[44].

Reported TFV resistance was a rare event, therefore the remaining studies reporting resistance (2/20) were investigated as potential outliers. On qualitative analysis both studies were small (n=10,11) with prior adefovir +/- lamivudine exposure. A cohort study in the USA performed between 1999-2007 [54], classified 1/10 individuals as having genotypic resistance to TFV. The individual had received prior treatment with adefovir and lamivudine and had adefovir resistance at baseline (A181V on direct sequencing and N236T on clonal analysis) which persisted during

TDF treatment (accepted TFV RAMs, Supplementary table 2A). In a retrospective study performed in 2012 in Italy[16], 1/11 individuals were classified as having genotypic resistance to TFV. This individual did not respond to prior lamivudine and adefovir treatment however did not have lamivudine or adefovir RAMs at baseline. They developed an A194T mutation on TFV (putative TFV RAM (Table 1B), Supplementary table 2B). Individuals with incomplete adherence were not excluded from any of these two studies.

In order to establish whether heterogeneity in the estimate for risk of TFV resistance could have been influenced by variations in thresholds for calling TFV RAMs, we compared the resistance criteria across papers (Supplementary figure 2C). Among the 26 papers included in the primary analysis for TFV (irrespective of whether previously naive or treatment experienced), only five classified mutations A181T/V and N236T known to be linked to reduced TFV sensitivity as RAMs (Supplementary table 2A, 2C), seven papers identified these mutations but did not classify them as resistant. Three of the 26 papers reported one or more of the putative mutations listed in Supplementary table 2B as RAMs, an additional 8 papers identified one or more of these combinations of mutations but did not report the individual as resistant.

**iii) Naive/Entecavir**

In previously NA-naive individuals prospectively followed up after starting ETV, heterogeneity was seen at years 2, 4 and ≥5 time points. None of the sensitivity analyses performed reduced the heterogeneity (Supplementary table 3B).

On qualitative analysis of heterogeneity of studies at year 2, the highest resistance risk estimate at 3.3% was a multi-centre clinical trial of HBeAg positive children, all individuals developed M204M/V, L180M, S202G mutations (the only other study recruiting children [49] was also in the same subgroup with a resistance risk of 0.0%). At years 4 and ≥5, both papers reporting no resistance used more stringent criteria for sequencing, only investigating individuals meeting the criteria for VBT, rather than all those with detectable viraemia. This may lead to an under-estimation of primary non-response to ETV in naive individuals. A retrospective cohort study in Korea[33] reported the highest risk of 3% at ≥5 years. The duration of follow up was longer here than the other trials in the same time bracket (101 vs 60-70 months) which may explain the higher proportion of resistance observed (however the same study also reported on previously NA experienced individuals, and a relatively low risk of resistance was identified).

iv) **Experienced/Entecavir**

In previously NA experienced individuals on ETV, heterogeneity was seen at 2, 3 and 5+ year timepoints. Sensitivity analysis only showed a reduction in heterogeneity when restricting based on sequencing criteria at year 2 (reducing heterogeneity from 97% to 62%) and presence of baseline RAMs at year 5, reducing heterogeneity from 93% to 80%. Supplementary figure 3 shows the impact of baseline RAMs across all papers, those studies excluding RAMs reported a lower risk of ETV resistance (<10%) than those where some or all individuals had RAMs at baseline (up to 51.5% resistance).

On qualitative analysis of the highest/lowest risk at each time point, at 2 years, the study with the lowest resistance risk, a clinical trial in Korea (2.9% resistance) excluded those with baseline RAMs but sequenced all individuals with detectable VL[24]. The study with the highest estimate of 36.4% studied individuals all of whom had genotypic resistance to lamivudine at baseline and were also refractory to adefovir[43]. The difference in baseline populations may contribute to heterogeneity here due to the influence of cross-resistance (Table 2).

At 3 years, the lowest estimate of 8.9% was a retrospective study of 146 individuals without baseline mutations[13]. The highest estimate (38.5%) came from a retrospective cohort of 13 individuals with detectable VL on treatment[25]. Details of prior NA exposure was not reported for this subgroup and baseline RAMs were not reported; only those with VBT were sequenced, however a line probe assay was used in addition to sequencing to determine genotypic resistance, and it is not clear whether the results presented are from sequencing alone, or the line probe assay.

The lowest estimate of resistance risk at ≥5 years was a retrospective cohort study in Korea[33] (3.6% ETV resistance), which excluded individuals with any RAMs at baseline and only performed sequencing on individuals with VBT. In contrast, in the study with the highest estimate (51.5% ETV resistance) all individuals had RAMs to other NAs at baseline and on follow up sequenced all individuals with a detectable viral load[56].

Four studies [10,33,52,58] used a lower dose of ETV (0.5mg OD) than the 1.0mg dose currently licensed for cases of resistance (where studies used multiple doses, data from the 1mg OD group was extracted). However it was not possible to assess the impact of dosing on heterogeneity as the majority of these studies were in a year group where no heterogeneity was observed. All studies except one[58] included individuals with prior LAM experience, so it was not possible to determine whether prior LAM (vs. other NA) was a particular risk factor for ETV resistance.

25. Hongthanakorn C, Chotiyaputta W, Oberhelman K, et al. Virological breakthrough and resistance in patients with chronic hepatitis B receiving nucleos(t)ide analogues in clinical

practice. *Hepatology*. 2011;53(6):1854-1863.
